# AI-Based Multi-Year Prediction of Incident Dementia in Patients with Atrial Fibrillation

**DOI:** 10.64898/2026.11.23.26368531

**Authors:** Sabyasachi Bandyopadhyay, Viren Srivastava, Rayan Ansari, Esteban Peralta, Prasanth Ganesan, Jake Stringer, Charles Sillet, Jingwen Huang, Chad Robichaux, Samuel Ruiperez-Campillo, Brototo Deb, Hui Ju Chang, Faisal M. Merchant, Paul Clopton, Chad R. Brodt, Albert J. Rogers, Neal K. Bhatia, Kyan Younes, Maarten Lansberg, Sanjiv M. Narayan

## Abstract

The burden of dementia is rising rapidly, but preventive strategies remain limited and few tools exist to identify individuals at risk *a priori*. This unmet need is especially relevant in atrial fibrillation (AF), a rapidly growing population at increased risk of dementia. We present MIND-AF, an AI-based model to predict 3 to 5-year risk of dementia from readily available bedside data. Developed in 108,672 AF patients without prior dementia within an academic–community network in California, MIND-AF predicted incident dementia in a held-out test cohort (n=32,238) with a C-index of 0.82 over 3.8 ± 4.6 years after AF diagnosis, outperforming age and clinical risk scores (CHA₂DS₂-VASc, HATCH; all p<0.001). MIND-AF enabled *a priori* stratification into low, medium and high-risk groups, with dementia incidence of 0.34%, 1.59% and 6.3% over 3 years and 0.7%, 3.4% and 13.0% over 5 years from AF diagnosis, respectively. MIND-AF provided similar performance in a sealed external population of 58,826 demographically distinct patients in Georgia (C-index 0.80), as well as pre-specified cohorts without neurodegenerative disease, mild cognitive impairment, stroke or transient ischemic attack. MIND-AF, freely available as an online web application, provides scalable point-of-care risk stratification for incident dementia in patients diagnosed with AF, is robust to baseline neurocognitive and cerebrovascular baseline states, and provides a bedside framework for intensified dementia prevention.

## Introduction

Cognitive decline is a leading cause of disability in ageing populations, and many populations have heightened risk. Atrial fibrillation (AF) affects over 60 million individuals worldwide ^1,2^, with rising prevalence due to screening efforts and the widespread use of wearable devices. Patients with AF are at high risk for dementia, for which there is increasing focus on early detection. While stroke prevention in AF is guided by validated risk stratification tools, no equivalent framework exists to identify individuals at highest risk of dementia. This gap is significant as stroke is an established risk factor for dementia, operating through mechanisms such as direct disruption of critical brain networks and triggering of neurodegenerative cascades post-stroke^3^. Contemporary guidelines for patients with AF increasingly recommend specific strategies in patients at elevated risk of dementia^2^, yet their implementation is limited by the paucity of tools to quantify individual dementia risk.

Dementia risk scores developed for general populations such as CAIDE (Cardiovascular Risk Factors, Aging, and Incidence of Dementia)^4^, BDSI (Brief Dementia Screening Indicator)^5^, ANU-ADRI (ANU Alzheimer’s Disease Risk Index)^6^ and DRS (Dementia Risk Score)^7^ show limited predictive value beyond age alone^8–10^, and may not capture the heightening of risk after the onset of predisposing conditions such as AF^11^. In populations with AF, risk scores for stroke such as CHA₂DS₂-VASc, APPLE and GARFIELD-AF^2,12^ modestly predict dementia beyond age at AF onset ^1,13^. Novel biomarkers can identify pathology for Alzheimer’s Disease (AD) before the onset of dementia, including amyloid in plasma and cerebrospinal fluid, or positron emission tomography (PET) imaging using a radiotracer that binds to aggregated tau protein^14^, yet these specialized tests are less suited for bedside screening.

We present MIND-AF (<u>M</u>odeling Incident <u>N</u>euro <u>D</u>egeneration in <u>A</u>trial <u>F</u>ibrillation), an AI-based system designed to quantify the risk of incident dementia and estimate time-to-event risk at AF diagnosis from readily available clinical features. MIND-AF, developed in a registry of 108,672 patients and validated in an independent, geographically-distinct cohort of 58,826 patients, stratifies individuals into three risk groups with markedly different time-dependent 3 year and 5 year outcomes (Fig. 1). MIND-AF maintained performance in pre-specified sub-populations with stringent exclusions of baseline cognitive impairment or cerebrovascular disease (stroke or transient ischemia attack), and is deployed as a web-based calculator for point-of-care use.

**Figure 1:**
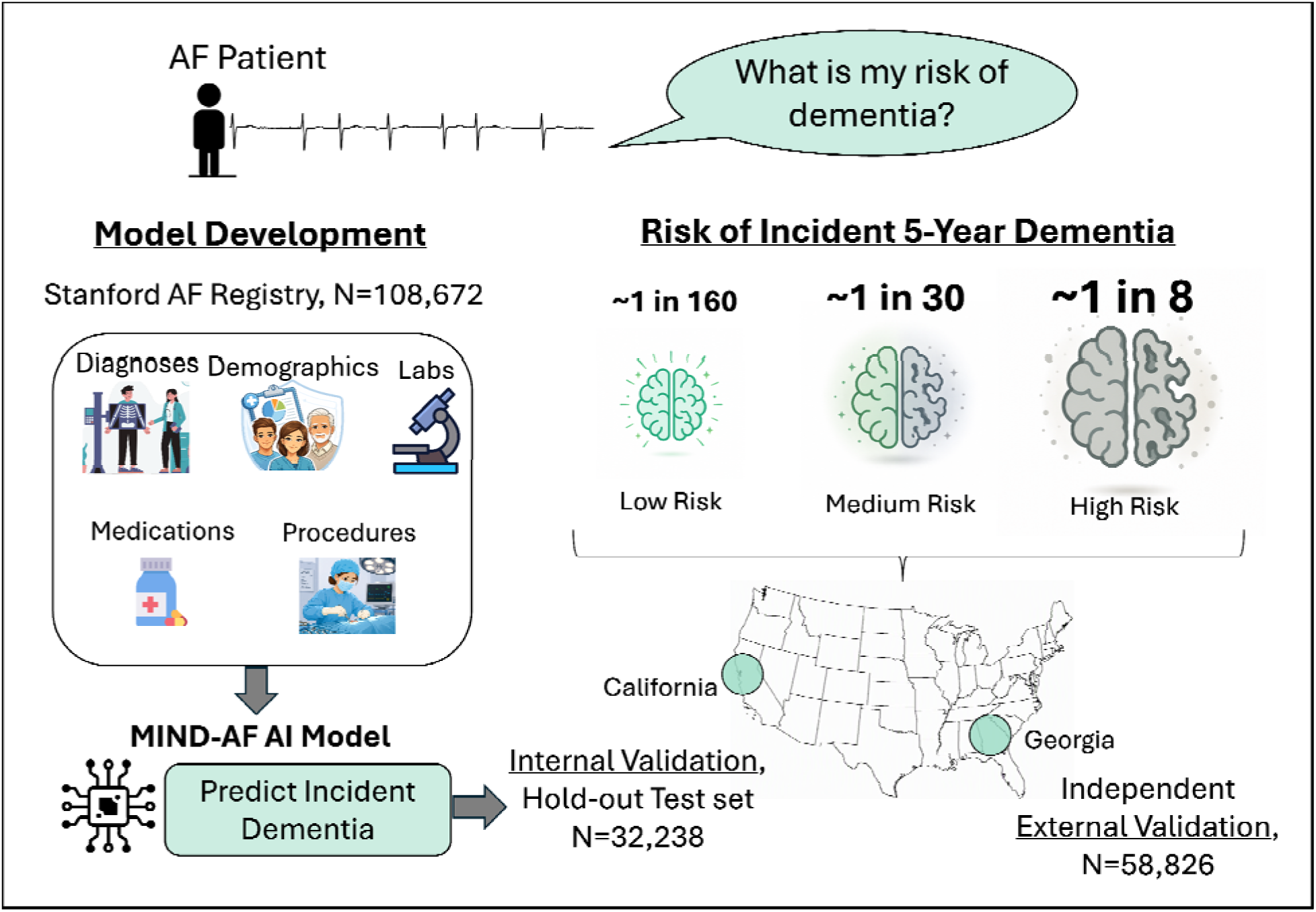
Conceptual workflow for MIND-AF (Modeling Incident Neurodegeneration in AF). Multimodal clinical and demographic data from patients with atrial fibrillation (AF) were used to develop MIND-AF. In an unseen hold-out cohort in California and a sealed independent validation cohort from Georgia, MIND-AF predicted incident AF better than existing risk scores and predicted time-dependent risk in three risk categories with significantly different 3 year and 5 year dementia incidence.

## Results

### Study population

We identified 108,672 patients followed at Stanford Healthcare in Northern California between 2000-2024 with ICD9 ode 427.X and ICD10 code I48.X indicating AF. We excluded patients with only atrial flutter, prior dementia (defined in Supplementary Table 1), those with dementia diagnosed within 1 year after AF diagnosis that could be overlooked at baseline, and those without longitudinal follow-up (fewer than two notes over > 1 year). Fig. 2 outlines the consort diagram resulting in our primary population of 96,688 individuals. For external validation, we curated a cohort of 80,571 patients with identical inclusion and exclusion criteria at Emory University in Atlanta, Georgia, of whom 58,826 served as the external validation test cohort (Extended Data Fig 1). Patients in Georgia exhibited several distinct characteristics, including higher average BMI and a greater proportion of African American patients, than the California population (Table 1).

**Figure 2:**
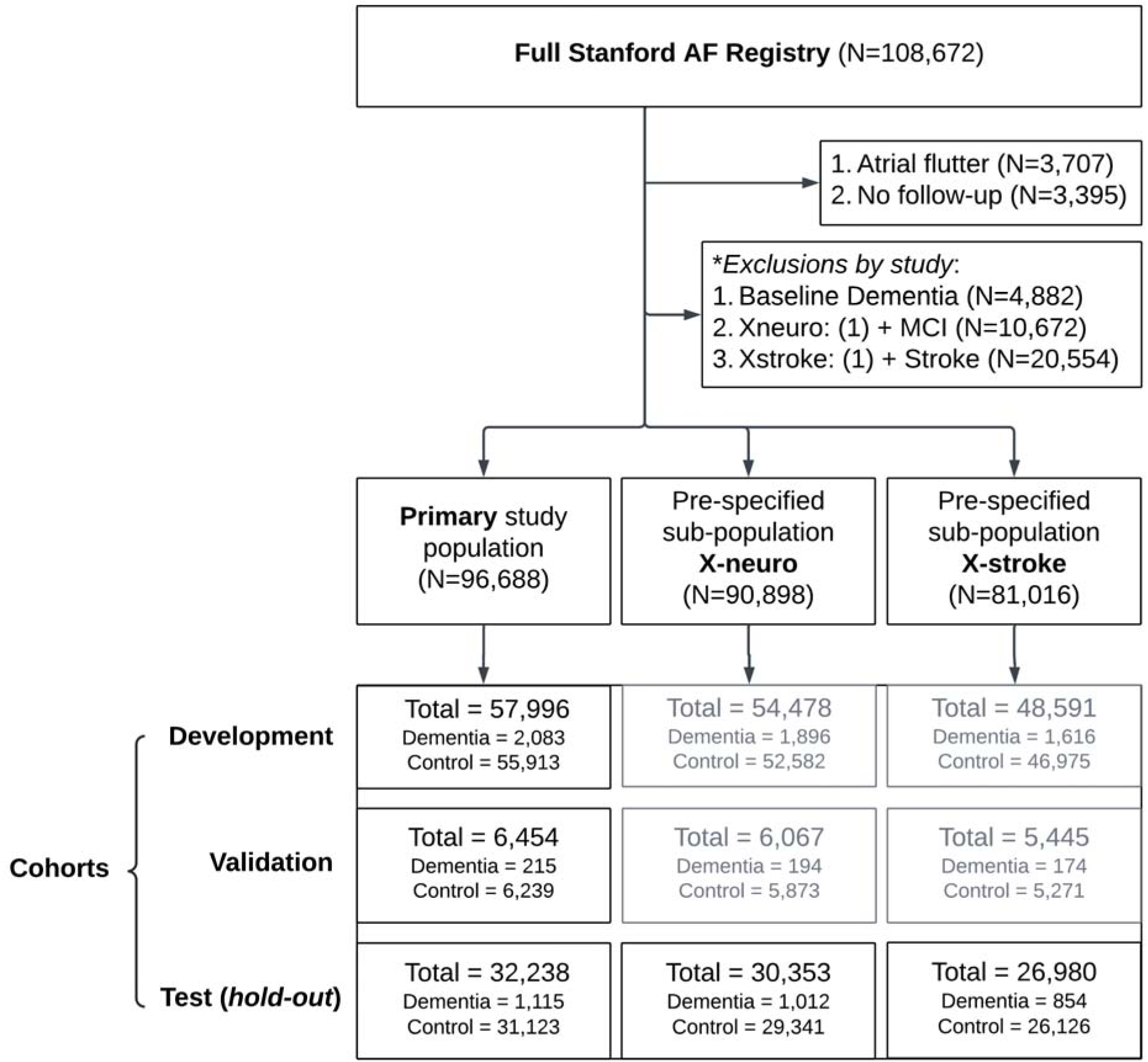
Consort Diagram for MIND-AF. The primary population for MIND-AF were patients with an AF diagnosis without baseline dementia. Pre-specified populations lacked any baseline cognitive impairment or dementia (X-neuro cohort), and lacked symptomatic TIA or stroke (X-stroke cohort).

**Table 1:** Characteristics of primary Stanford cohort and external Emory test cohort.

| **Feature** | **Development set (Stanford)** |  |  | **Hold-out test set (Stanford)** |  |  | **External test set (Emory)** |  |  |
|---|---|---|---|---|---|---|---|---|---|
|  | **No dementia (n = 55,913)** | **Dementia (n = 2,083)** | **P value** | **No dementia (n = 31,123)** | **Dementia (n = 1,115)** | **P value** | **No dementia (n = 56,434)** | **Dementia (n = 2,392)** | **P value** |
| **Demographics** |  |  |  |  |  |  |  |  |  |
| AF diagnosis age, years | 70.2 ± 14.0 | 79.1 ± 9.1 | **<0.001** | 70.2 ± 14.1 | 79.2 ± 9.0 | **<0.001** | 67.4 ± 13.3 | 75.3 ± 9.8 | **<0.001** |
| BMI, kg/m<sup>2</sup> | 27.3 ± 5.5 | 25.0 ± 5.0 | **<0.001** | 27.3 ± 5.5 | 25.1 ± 5.0 | **<0.001** | 28.7 ± 5.5 | 28.2 ± 5.5 | **<0.001** |
| Female sex, n (%) | 23,266 (41.6) | 1,130 (54.2) | **<0.001** | 13,046 (41.9) | 574 (51.5) | **<0.001** | 23,839 (42.2) | 1,231 (51.5) | **<0.001** |
| **Race** |  |  |  |  |  |  |  |  |  |
| White, n (%) | 36,201 (64.7) | 1,512 (72.6) | **<0.001** | 20,135 (64.7) | 788 (70.7) | **<0.001** | 34,229 (60.7) | 1,447 (60.5) | **<0.001** |
| **Cardiovascular comorbidities** |  |  |  |  |  |  |  |  |  |
| Congestive heart failure, n (%) | 11,679 (20.9) | 366 (17.6) | **<0.001** | 6,584 (21.2) | 216 (19.4) | 0.15 | 10,084 (17.9) | 230 (9.6) | **<0.001** |
| Hypertension, n (%) | 30,615 (54.8) | 1,326 (63.7) | **<0.001** | 16,917 (54.4) | 716 (64.2) | **<0.001** | 26,293 (46.6) | 899 (37.6) | **<0.001** |
| Hyperlipidemia, n (%) | 22,588 (40.4) | 939 (45.1) | **<0.001** | 12,491 (40.1) | 517 (46.4) | **<0.001** | 14,817 (26.3) | 381 (15.9) | **<0.001** |
| Stroke or TIA, n (%) | 6,887 (12.3) | 355 (17.0) | **<0.001** | 3,811 (12.2) | 192 (17.2) | **<0.001** | 5,747 (10.2) | 259 (10.8) | 0.325 |
| Valvular surgery, n (%) | 1,561 (2.8) | 38 (1.8) | **0.008** | 858 (2.8) | 22 (2.0) | 0.115 | 297 (0.5) | 3 (0.1) | 0.011 |
| Catheter ablation for AF, n (%) | 117 (0.2) | 0 (0.0) | 0.037 | 68 (0.2) | 0 (0.0) | 0.118 | 1 (0.0) | 0 (0.0) | 1.0 |
| **Non-cardiovascular comorbidities** |  |  |  |  |  |  |  |  |  |
| Diabetes mellitus, n (%) | 10,076 (18.0) | 407 (19.5) | 0.077 | 5,677 (18.2) | 216 (19.4) | 0.337 | 10,351 (18.3) | 279 (11.7) | **<0.001** |
| Depression, n (%) | 4,812 (8.6) | 278 (13.3) | **<0.001** | 2,767 (8.9) | 145 (13.0) | **<0.001** | 3,472 (6.2) | 92 (3.8) | **<0.001** |
| Parkinson's disease, n (%) | 448 (0.8) | 49 (2.4) | **<0.001** | 243 (0.8) | 32 (2.9) | **<0.001** | 261 (0.5) | 45 (1.9) | **<0.001** |
| Schizophrenia, n (%) | 401 (0.7) | 29 (1.4) | **<0.001** | 243 (0.8) | 10 (0.9) | 0.666 | 216 (0.4) | 7 (0.3) | 0.594 |
| Mild cognitive impairment, n (%) | 290 (0.5) | 33 (1.6) | **<0.001** | 143 (0.5) | 28 (2.5) | **<0.001** | 173 (0.3) | 23 (1.0) | **<0.001** |
| Brain injury, n (%) | 533 (1.0) | 34 (1.6) | **0.002** | 284 (0.9) | 13 (1.2) | 0.384 | 414 (0.7) | 17 (0.7) | 0.995 |
| **Medications** |  |  |  |  |  |  |  |  |  |
| Coumadin/Warfarin, n (%) | 3,942 (7.1) | 176 (8.4) | 0.015 | 2,253 (7.2) | 62 (5.6) | 0.033 | 1,917 (3.4) | 72 (3.0) | 0.333 |
| HMG-CoA reductase inhibitors, n (%) | 17,752 (31.7) | 548 (26.3) | **<0.001** | 9,774 (31.4) | 293 (26.3) | **<0.001** | 21,757 (38.6) | 649 (27.1) | **<0.001** |

Abbreviations. BMI; Body Mass Index, MIND-AF; Modeling Incident Neurodegeneration in Atrial fibrillation, HMG-CoA, hydroxymethylglutaryl-coenzyme A, TIA; Transient Ischemic Attack. \*p-value < 0.01.

In the primary (California) cohort, 3,413 patients developed dementia (3.53%) an average of 3.8 ± 4.6 years after AF diagnosis. The age-dependent incidence of dementia rose from 0.9% in patients diagnosed with AF at < 65 years to 7% in those diagnosed with AF at ≥ 85 years (Supplementary Fig 1).

We partitioned the population randomly into development, validation and held-out test cohorts as summarized in Table 1, with extended characteristics in Supplementary Table 2, and ICD codes for comorbidities in Supplementary Table 3. To test model generalizability, we identified a pre-specified X-neuro sub-population with exclusion of neurodegenerative diseases and mild cognitive impairment (MCI) at baseline, and an X-stroke subpopulation with exclusion of stroke and TIA at baseline (Online Methods; Fig 2).

### Prediction of Incident Dementia

In developing MIND-AF, we trained competing architectures (XGBoost, GLMBoost, Cox Proportional Hazard (Cox PH) and Deep survival models) in the Stanford Development cohort, of which XGBoost provided the highest risk predictive accuracy in the validation cohort (Extended Data Table 1) and was selected as the final architecture.

In the held-out California test cohort, which represents a real-world screening population retaining pre-existing MCI, non-dementia neurodegenerative diseases, stroke and TIA, MIND-AF achieved a C-index of 0.82 (95% CI 0.81-0.83) and AUROC of 0.85 (95% CI 0.84-0.87) for dementia within 5-years of AF diagnosis (Table 2). We also evaluated two prespecified subcohorts (Table 2). In X-neuro, which excludes baseline MCI and non-dementia neurodegenerative diagnoses, the C-index was 0.81 (95% CI 0.80–0.82) and 5-year AUROC was 0.84 (95% CI 0.82–0.85). In X-stroke, which excludes baseline stroke and TIA, the C-index was 0.83 (95% CI 0.81–0.84) and 5-year AUROC was 0.85 (95% CI 0.84–0.87).

**Table 2:** Performance of MIND-AF and clinical scores in predicting incident dementia.

|  | **Time-to-dementia** |  |  | **Landmark Analysis (dementia within 5-years)** |  |  |  |  |
| **Model** | **C-index (95% CI)** | **Hazard ratio low vs high (95% CI)** | **Hazard ratio per 1-SD (95% CI)** | **AUROC (95% CI)** | **Sensitivity (95% CI)** | **Specificity (95% CI)** | **Precision (95% CI)** | **F1-score (95% CI)** |
|---|---|---|---|---|---|---|---|---|
| **Primary Cohort (no baseline dementia)** |  |  |  |  |  |  |  |  |
| **MIND-AF** | 0.82 (0.81–0.83) | 16.96 (14.02–20.53) | 4.24 (3.93-4.58) | 0.85 (0.84–0.87) | 0.72 (0.69–0.76) | 0.81 (0.80–0.82) | 0.18 (0.17–0.20) | 0.29 (0.27–0.32) |
| *CHA<sub>2</sub>DS<sub>2</sub>-VASc* | 0.73 (0.72–0.75) | — | 2.24 (2.12-2.36) | 0.76 (0.74–0.78) | 0.97 (0.95–0.98) | 0.31 (0.30–0.32) | 0.07 (0.07–0.08) | 0.14 (0.13–0.15) |
| *HATCH* | 0.69 (0.67–0.71) | — | 1.83 (1.73-1.93) | 0.72 (0.70–0.73) | 0.72 (0.68–0.75) | 0.61 (0.60–0.62) | 0.10 (0.09–0.11) | 0.17 (0.16–0.19) |
| **Pre-Specified X-Neuro Cohort (no baseline dementia or mild cognitive impairment)** |  |  |  |  |  |  |  |  |
| **MIND-AF** | 0.81 (0.80–0.82) | 14.27 (11.90–17.11) | 4.02 (3.73-4.33) | 0.84 (0.82–0.85) | 0.74 (0.70–0.77) | 0.77 (0.76–0.78) | 0.17 (0.16–0.18) | 0.28 (0.26–0.30) |
| *CHA<sub>2</sub>DS<sub>2</sub>-VASc* | 0.72 (0.71–0.74) | — | 2.17 (2.06-2.29) | 0.76 (0.74–0.78) | 0.95 (0.93–0.97) | 0.35 (0.34–0.36) | 0.07 (0.06–0.08) | 0.13 (0.11–0.14) |
| *HATCH* | 0.68 (0.66–0.70) | — | 1.80 (1.70-1.89) | 0.71 (0.69–0.73) | 0.65 (0.60–0.69) | 0.67 (0.66–0.68) | 0.09 (0.08–0.10) | 0.16 (0.14–0.17) |
| **Pre-specified X-Stroke Cohort (no baseline dementia, Stroke or TIA)** |  |  |  |  |  |  |  |  |
| **MIND-AF** | 0.83 (0.81–0.84) | 18.26 (14.77–22.57) | 4.63 (4.22-5.07) | 0.85 (0.84–0.87) | 0.71 (0.66–0.75) | 0.83 (0.82–0.84) | 0.17 (0.16–0.19) | 0.28 (0.25–0.30) |
| *CHA<sub>2</sub>DS<sub>2</sub>-VASc* | 0.74 (0.72–0.76) | — | 2.40 (2.24-2.57) | 0.75 (0.73–0.76) | 0.95 (0.93–0.97) | 0.31 (0.30–0.32) | 0.08 (0.07–0.08) | 0.15 (0.14–0.16) |
| *HATCH* | 0.69 (0.67–0.71) | — | 1.83 (1.72-1.95) | 0.71 (0.69–0.73) | 0.71 (0.67–0.75) | 0.61 (0.60–0.62) | 0.10 (0.09–0.11) | 0.18 (0.16–0.19) |

Abbreviations. AF; Atrial fibrillation, AUROC; Area under receiver operating curve, C-index; Concordance index, MIND-AF; Modeling incident neurodegeneration in atrial fibrillation.
Footnote. Operating-point metrics (sensitivity, specificity, precision, F1) were computed at pre-specified thresholds — MIND-AF at 5.2% predicted 5-year risk, CHA<sub>2</sub>DS<sub>2</sub>-VASc at ≥2, and HATCH at ≥2 — rather than data-derived cutpoints; AUROC is threshold-independent. For the landmark analysis, patients censored before the 5-year horizon were excluded.

MIND-AF predicted incident dementia significantly better than age, the most effective current predictor of dementia ^8–10^ and well-studied clinical risk scores (Table 2). Net reclassification improvement (NRI) was +19.2% (95% CI 15.7-22.8) against age and +42.3% (95% CI 38.4-46.0) against CHA₂DS₂-VASc. The integrated discrimination index (IDI) also improved (+3.6% compared to age; +6.9% compared to CHA₂DS₂-VASc), reflecting substantially greater separation between dementia and non-dementia risk distributions. Figure 3 shows a multivariable Cox model including both MIND-AF score and age at AF diagnosis, in which each 1-s.d. increase in the MIND-AF score was associated with a significantly greater increase in the hazard for dementia (HR 3.52 (95% CI 3.08-4.04) than a 1-s.d. increase in age (HR 1.32 (95% CI 1.12-1.55), p<0.001). Across age strata, MIND-AF attributed higher risk than age alone, with an age-weighted global relative risk ratio of 1.50 (95% CI 1.47–1.53; Fig. 3).

**Figure 3:**
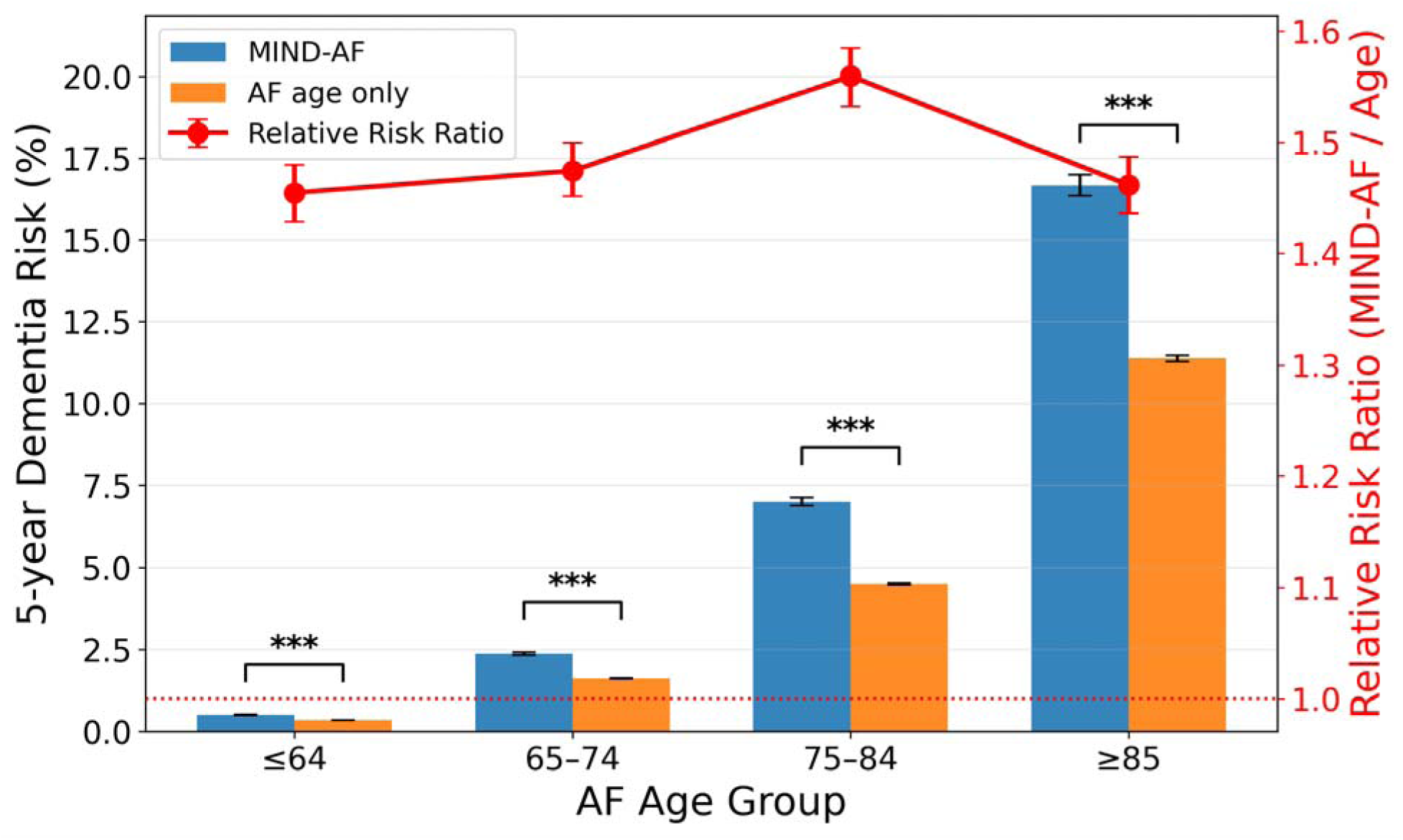
MIND-AF identifies dementia risk beyond age at AF diagnosis. Predicted 5-year dementia risk across AF age groups using MIND-AF versus age at AF diagnosis alone. Bars show absolute predicted risk with 95% confidence intervals. The red line shows the relative risk ratio for MIND-AF versus age alone, with 95% confidence intervals. MIND-AF assigned higher risk than age alone across all age strata, indicating incremental risk discrimination beyond age. ***P<0.001.

### Time-Dependent Prediction of Dementia Risk

Determining risk over time is particularly important as dementia typically evolves over years. We defined meaningful risk thresholds for developing dementia within 5 years of AF diagnosis using previously reported decision curve analysis (DCA)^15^ (Supplementary Fig. 2). This identified low, intermediate- and high-risk groups with incidence of ≤2%, >2 to 5.2%, and >5.2%, respectively.

Figure 4 shows that MIND-AF yielded a 5-year dementia incidence of 0.7%, 3.4% and 13.0% in low, intermediate and high risk-strata of the held-out California test cohort, with 3-year incidences of 0.34%, 1.59% and 6.3%, respectively. Observed event rates therefore separated well across the prespecified risk strata, with a 16.9-fold higher 5-year hazard of dementia in the high-versus low-risk group (HR 16.9, 95% CI 14.0–20.5). Predicted and observed risks were well calibrated across risk groups (Extended Data Fig. 2).

**Figure 4:**
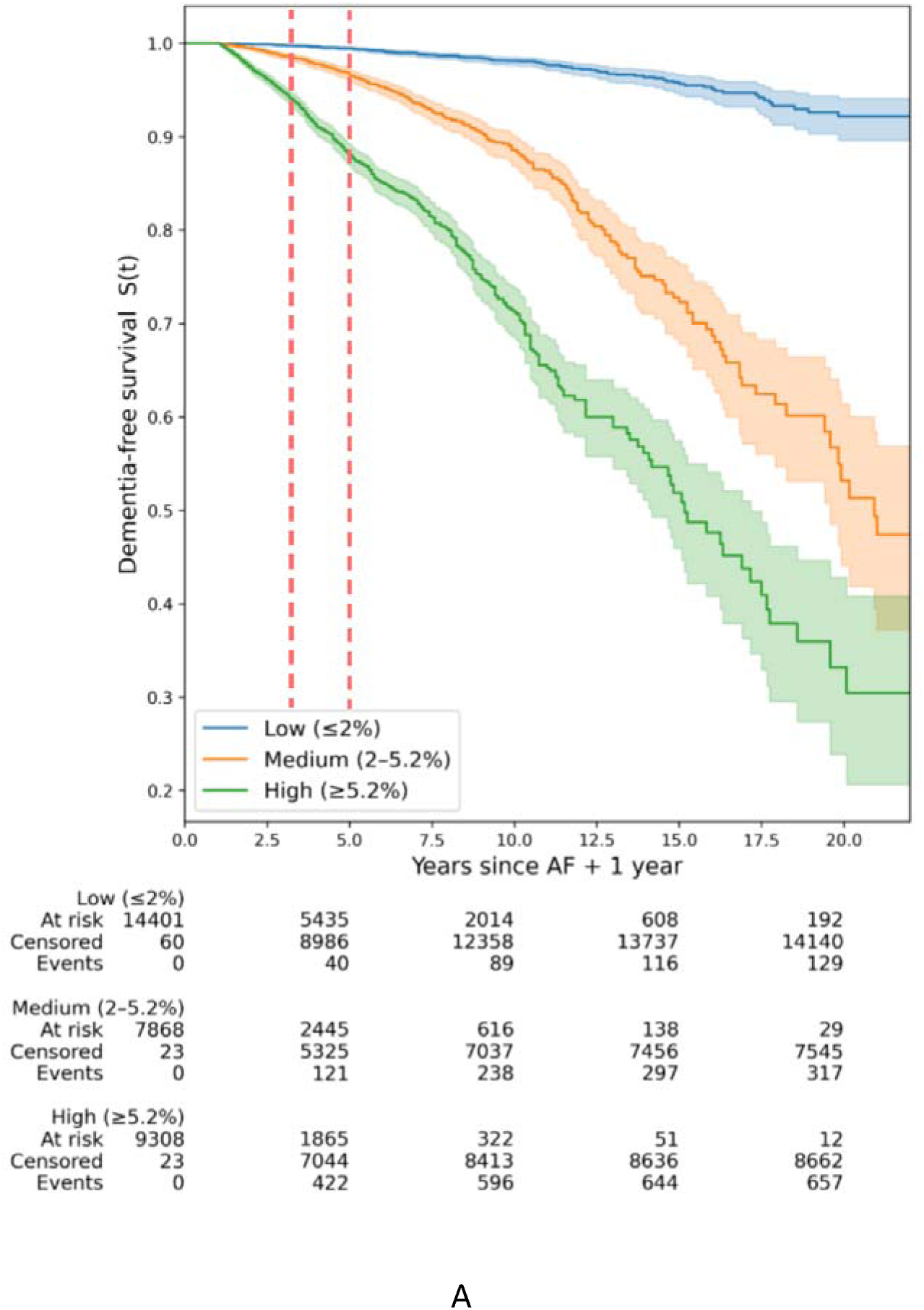
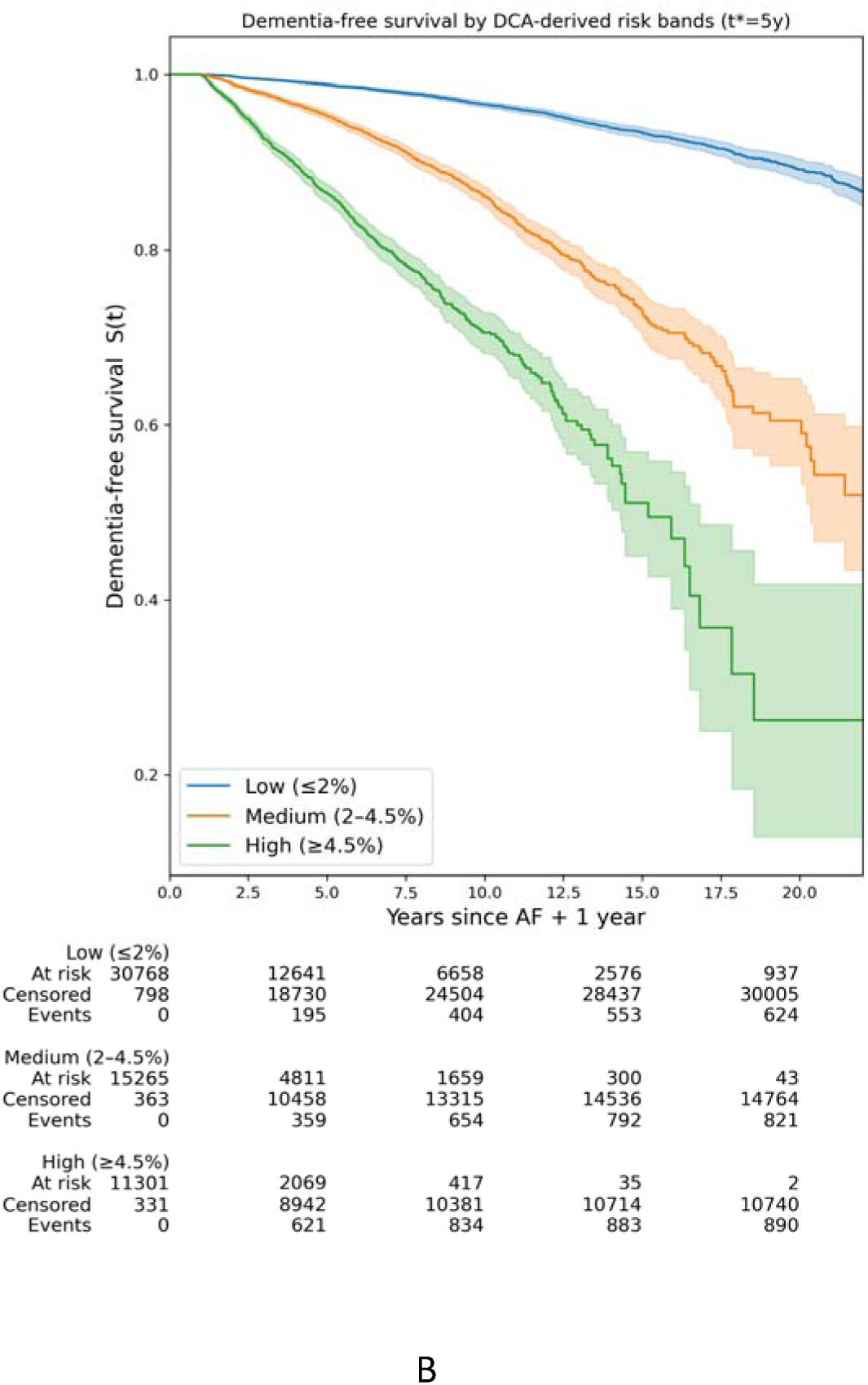
MIND-AF Risk Strata separate Dementia-free survival in the California test cohort and Georgia external validation cohort. Kaplan–Meier curves show dementia-free survival across decision curve analysis–derived 5-year predicted risk groups. Red dotted lines mark the 3-year and 5-year time points after AF diagnosis. **(a)** Stanford hold-out test cohort (n = 32,238) stratified as low, intermediate and high. **(b)** Emory external validation cohort (n = 58,826) stratified using the same predicted-risk framework. Thresholds were fixed at low (≤2%), intermediate (>2–4.5%) and high (≥4.5%), derived from the web-deployed 22-feature model. In both cohorts, the three risk groups show clear, progressive separation sustained throughout follow-up, demonstrating preserved risk stratification on external validation. Shaded areas indicate 95% confidence intervals. The numbers at risk, censored observations, and incident dementia events are shown below each time point. Patients were censored at last follow-up if dementia did not occur.

Time-dependent risk stratification was maintained in the prespecified X-neuro subcohort (high-versus-low hazard ratio 14.3, 95% CI 11.9 - 17.1) and in the X-stroke subcohort (18.3, 95% CI 14.8 - 22.6; Table 2), with comparable risk bands (Supplementary Table 4). Thus, MIND-AF provided robust time-dependent risk stratification across populations free of baseline dementia and broadly defined cognitive impairment or cerebrovascular disease.

### External Validation in Georgia Cohort

MIND-AF successfully stratified patients in the external Georgia cohort into low, intermediate and high-risk groups with 5-year dementia incidence of 1.1%, 4.6%, and 13.5%, respectively (Fig 4b), closely replicating the separation observed in the primary cohort. MIND-AF demonstrated a time-to-event C-index of 0.79 (95% CI 0.78–0.80), and AUROC at 5 years of 0.83 (95% CI 0.82-0.84) for classifying dementia that outperformed clinical risk scores (Extended Data Table 2), similar to the California cohort. The performance of MIND-AF in the external Georgia population without site-specific retraining suggests that it encodes features shared across clinically and demographically distinct populations.

### Clinical Features Explaining Model Predictions

We probed clinical features in MIND-AF that contribute most to its performance using Shapley analysis of the final XGBoost model (Extended Data Fig 3a and Supplementary Table 5). The separation of low from intermediate, and intermediate from high risk groups were explained primarily by age followed by a multidomain combination of cardiometabolic, sociodemographic cardiometabolic, neurocognitive and cerebrovascular features. The contribution of features was similar for the primary California and external Georgia validation populations (Extended Data Fig 4). The relative contribution of age diminished in the higher risk strata, indicating the multidomain pathology of dementia which agrees with several longitudinal reports on dementia risk factors^16^.

### Clinical translation: web-based risk calculator

For point-of-care use, MIND-AF was translated into a web-based estimator of individualized 5-year dementia risk using a reduced set of 22 features derived by Bayesian information criterion–guided ablation (Extended Data Fig 5). The app preserved discrimination in relation to the full model and, in the internal held-out test cohort, provided a C-index of 0.821 (Supplementary Fig. 3). If some input data are missing, the web-app implements a feature-specific fall back to median values in the training set (Online Methods), and preserves patient privacy^17^ by processing inputs locally without storing user data. This tool is freely available at https://github.com/NarayanLab/ml-dementia-prediction.

## Discussion

We report an AI-based approach to predict incident dementia in patients with atrial fibrillation (AF) from their time of diagnosis, that outperforms the widely used predictor of age and several clinical risk scores. The AI-system, MIND-AF, developed in 64,450 patients in California, provided a hazards ratio exceeding 16 in separating individuals at high from low risk for developing dementia over a period of 5 years from AF diagnosis in 32,238 patients in California and a sealed external cohort of 58,826 patients in Georgia. MIND-AF was robust to baseline comorbidities and performed similarly well in prespecified subpopulations lacking baseline neurodegenerative diseases, cognitive impairment, or cerebrovascular disease. MIND-AF was designed to minimize the need for specialized biometric or imaging data, and explainability analysis showed that MIND-AF encoded several potentially modifiable features for dementia ^16^.

By enabling bedside stratification of elevated risk for dementia in patients with AF, MIND-AF provides an actionable bedside tool to enable expedited referral for neurocognitive evaluation and to identify individuals at particularly high risk in whom novel preventative therapies could be applied.^2,18–20^. MIND-AF is freely available as an online web calculator to facilitate such use.

### Clinical value

AI and agentic systems have recently been reported to predict dementia, although some require sophisticated multiomic markers or imaging that are not universally available ^21^. MIND-AF, conversely, was developed using readily available bedside clinical data, including clinical reports of laboratory and imaging data, that may contribute to its generalizability. MIND-AF was trained using a blanking period of 1 year, which may minimize the inclusion of individuals with subclinical or impending dementia at baseline and which may have contributed to its robust performance. The online web-application is designed for point of care use and allows users to generate risk estimates even when some features are not available using in-built feature-specific fallbacks to Stanford development set median values (Online methods).

In the general population, a 2024 Lancet Commission identified hypertension, diabetes, obesity and depression among 14 potentially modifiable risk factors for dementia, alongside physical inactivity, social isolation and high LDL cholesterol ^16^. Interpretability analysis showed that many contributory features in MIND-AF overlap these modifiable risk factors (Extended Data Fig. 3)^22^, supporting the potential use of this AI-approach in early referral for potential preventive strategies. AF may accelerate risk of cognitive decline through distinct pathways^23,24^, including the propensity of AF patients to stroke/TIA.

### Future applications

Extension of this AI-approach may define risk across still-broader populations, such as identifying at risk populations for specific clinical trials of preventative strategies. Several strategies are being tested including lifestyle intervention^18^, anticoagulation even in patients below current thresholds to slow vascular dementia ^19,20^, or other approaches ^2^. Potential integration with an electronic health record could populate routinely recorded inputs automatically to estimate dementia risk at the time when AF diagnosis is entered into a patient’s chart. Broader work could extend this approach to other populations at heightened risk for dementia, for which this model could be used for pre-training to reduce the data burden for novel models.

### Limitations

Limitations of this study include its retrospective registry design, and so prospective trials to test the predictive value of MIND-AF are needed. Although the primary population from California and external validation population from Georgia differed in demographics, further validation datasets could be applied. Models were developed based on diagnoses from ICD codes, and future improvements could include natural language processing of free-text clinical neurological notes to better capture baseline features and detect incident events. Dementia etiology-specific endpoints were not analyzed separately, as subtype-level event incidence rates are too small to support stable time-to-event modelling and because administrative codes suboptimally discriminate dementia subtypes ^25^. Future studies incorporating neuroimaging such as MRI, amyloid, and tau PET scans, fluid biomarker such as plasma ptau217 and genetic data such as apoe genotype may further refine prediction and elucidate mechanisms^26^ at the potential cost of reducing applicability as a broad screening tool. Future studies could evaluate whether these imaging and fluid biomarkers complement MIND-AF risk stratification by characterizing disease biology and informing eligibility for mechanism-specific clinical trials. Broader proteomic signatures may provide additional information but currently lack established disease specificity in this setting.

## Online Methods

### Study population and definitions

The institutional review boards at Stanford University (protocol 55244) and Emory University (protocol 2023P005594) approved this study. We analyzed electronic health record data from 4,227,995 adults in the Stanford healthcare system (2000–2024). AF was identified using validated ICD-9 and ICD-10 codes, and in keeping with prior studies we excluded isolated atrial flutter ^27^ (Supplementary Table 1). We excluded patients with baseline dementia (including diagnoses within 1 year of AF onset) and those without follow-up, defined as fewer than 2 clinical notes beyond >1 year after AF diagnosis. To evaluate the sensitivity of dementia risk prediction to cohort definitions and robustness of MIND-AF, we constructed three cohorts differing in baseline neurocognitive and cerebrovascular exclusions.

These are as follows

1. Primary California cohort (MIND-AF) - We exclude baseline dementia and predict the risk of incident dementia. This definition reflects a real-world clinical setting where AF patients can have pre-existing cognitive impairment or stroke when diagnosed with AF. This resulted in a study population of 96,688 patients (Fig 2).
2. Neurodegenerative disease-free cohort (X-neuro)–We exclude patients with baseline neurodegenerative diseases or mild cognitive impairment (ICD codes G31.84, F06.7, G12.21–G12.25, G10, A81.0, A81.81, A81.82, A81.83, A81.9) and predict the risk of incident dementia. This resulted in a study population of 90,898 patients (Fig 2)
3. Cerebrovascular disease-free cohort (X-stroke)-We exclude patients with baseline dementia and stroke or transient ischemic attack (TIA) (ICD codes I60, I61, I62, I63, I64, I65, I66, G45, G46) and predict the risk of incident dementia. This definition was used to study dementia risk independent of stroke. It resulted in a study population of 81,016 patients (Fig 2)

The complete list of diseases and their ICD codes used to define the above cohorts is shown in Supplementary Table 1. Models developed across all cohorts had identical feature engineering, training, hyperparameter tuning, and evaluation.

### Data extraction and preprocessing

Clinical variables included demographics, comorbidities, procedures, medications and laboratory values. Socioeconomic status was estimated using area deprivation index^28^. Missing data were handled using median imputation with indicator variables. Values were restricted to the 1st–99th percentile range, and missing data were assigned a Boolean flag (indicator) variable for later identification.

### Model development

Data were partitioned into development, validation and test cohorts^29^ (Fig. 2). Cox proportional hazards models, fitted using penalized likelihood were used as a statistical baseline for the longitudinal risk modeling task. ML models for this task included XGBoost, DeepSurv^30^, and GLMBoost^31^. Hyperparameters were optimized using randomized search and based on the performance in the validation set. The final model was evaluated on the locked internal test cohort and applied to the external test cohort.

### Risk stratification and evaluation

Absolute 3-year and 5-year risk for dementia was estimated using Breslow’s method. Decision curve analysis^15^ was used to define thresholds for low, intermediate and high risk (Supplementary Fig. 2). Kaplan–Meier curves evaluated dementia-free survival differences in the test cohort. Performance metrics included C-index for risk modeling and AUROC for classification.

### Model interpretation and deployment

Feature contributions were assessed using SHAP. A reduced-feature model was derived using Bayesian information criterion–guided recursive feature addition (Supplementary Fig. 3). The final model was deployed as a web-based application (Extended Data Fig. 5). The web app uses a React frontend and a FastAPI backend. The model handles missing data during deployment by imputing with Stanford development set medians. The model uses only deidentified data, and does not store user data for its operation, thus preserving patient privacy.

### Statistical analysis

Continuous data are represented as mean ± standard deviation unless otherwise indicated. Categorical data are represented as sample size (percentage of population) unless otherwise indicated. For univariate analyses, categorical variables with greater than two classes were evaluated using the Chi squared test, while binary variables were evaluated using the two-sample Z-test for proportions. Continuous variables were compared using the two-sample T-test with unequal variance. Variables which satisfied p < 0.10 in univariate analyses were carried forward to multivariate modeling. Multivariate Cox proportional hazards testing was used to assess hazard ratios, 95% confidence intervals, and p-values for all variables.

Using the optimal XGBoost model, we used Breslow’s method to estimate baseline survival, computed an absolute 5-year dementia risk and performed DCA with inverse-probability of censoring weights across thresholds 0.02–0.30 to quantify net benefit versus blanket treatment strategies like “treat all” or “treat none”.

For a neural network model termed DeepSurv^30^, we estimated the best performing architecture and training regimen using a randomized Sobol search centered around the optimal hyperparameters found in the original study. More details of the hyperparameter tuning of this model can be found in the Supplementary Methods.

We conducted time-to-event analysis using the index time (t0) as the time of AF diagnosis. For each patient, follow-up time was calculated as (i) time to dementia diagnosis, or (ii) time to last known follow up (patients without dementia). A two-sided alpha-level of 0.05 was used to assign statistical significance for all tests.

## Data Availability

All data produced in the present study are available upon reasonable request to the authors

## Data Availability

Upon reasonable written request, subsets of data used for this study will be made available to academic researchers, but patient-level data from the development cohort are not publicly available due to privacy and institutional restrictions.

## Code Availability

The web-app is freely accessible through a public URL: https://github.com/NarayanLab/ml-dementia-prediction to facilitate point-of-care evaluation and external validation. All scripts required to reproduce the modeling workflow and deploy the calculator are provided.

## Acknowledgements

SB is supported by the American Heart Association Postdoctoral Research Fellowship (25POST1361932). A.J.R. is supported by the National Institutes of Health (K23 HL166977) and the American Heart Association Career Development Award (23CDA933663). K.Y. is supported by the National Institutes of Health/National Institute of Ageing (K23 AG09073301A1), the Alzheimer’s Association (AACSF-24-1307411), the Iqbal Farrukh and Asad Jamal Fund, and the Erb Family Foundation. This study was funded, in part, by grants from the National Institutes of Health NIH to S.M.N (R01 HL83359 The Dynamics of Human Atrial Fibrillation; HL149134, Machine Learning in Atrial Fibrillation).

## Author Contributions

SB. Conceptualization and design of project. Assisted in curation of data. Developed machine learning models, performed data analysis and assisted in web app development. Drafted the first version of the manuscript and edited all versions.

VS. Conceptualization and design of project. Assisted in curation of data. Performed data analysis. Assisted in drafting the first version of the manuscript.

RA. Performed data analysis and web app development.

EP. Assisted in curation of data and performed data analysis.

PG. Provided intellectual input. Assisted in data analysis. Edited figures and the final version of the manuscript.

JS. Performed data analysis. Assisted in drafting the first version of the manuscript.

CS. Assisted in drafting the first version of the manuscript and edited following versions.

JH. Assisted in curation of external validation data, and manuscript editing and review.

CR. Assisted in curation of external validation data, and manuscript editing and review..

SRC. Provided intellectual input. Edited the final version of the manuscript.

BD. Assisted in conceptualization and design of the project. Edited the final version of the manuscript.

HJC. Assisted in curation of data.

FMM. Provided intellectual input. Edited the manuscript.

PC. Assisted in statistical analyses. Reviewed and edited the manuscript.

CRB. Took part in conceptualization of the study and provided intellectual input.

AJR. Provided intellectual input throughout study performance. Edited manuscript.

NKB. Provided intellectual input, curated and supervised analysis in independent test cohort, Edited manuscript.

KY. Provided intellectual input, assisted in designing the ancillary models. Reviewed and edited versions of the manuscript.

ML. Provided intellectual input, assisted in designing the ancillary models. Assisted in designing the ancillary models. Reviewed and edited versions of the manuscript.

SMN. Conceptualization and design of the project. Reviewed all data analyses. Assisted in writing the first version of the manuscript, reviewed and edited all following versions. Responsible for obtaining funding.

## Competing Interests

SB. None related to this project. Intellectual property rights owned by the University of Florida.

SRC. reports equity, intellectual property, and consulting with Physcade Inc, and consulting with Stanford University.

PG. None related to this project. Intellectual property rights owned by the Florida Atlantic University.

PC. Consulting fees, PhysCade Inc.

CRB. Consulting relationships with Abbott, J&J Medtech, Boston Scientific and Atricure.

NKB reports consulting fees from Abbott.

AJR reports consulting fees and/or equity in WearLinq Inc, YorLabs Inc, Abbott, and Johnson & Johnson MedTech, and EBR Systems and intellectual property rights from Stanford University. KY reports consulting fees from Taudia Inc.

SMN. None related to this project. Intellectual property owned by UC Regents and Stanford University. Equity in Lifesignals.ai, PhysCade, Inc. Royalties from Uptodate Inc.

The following authors report no following competing interests. VS, RA, EP, JS, BD, HJC, ML.

## Extended Data

**Extended Data Figure 1.**
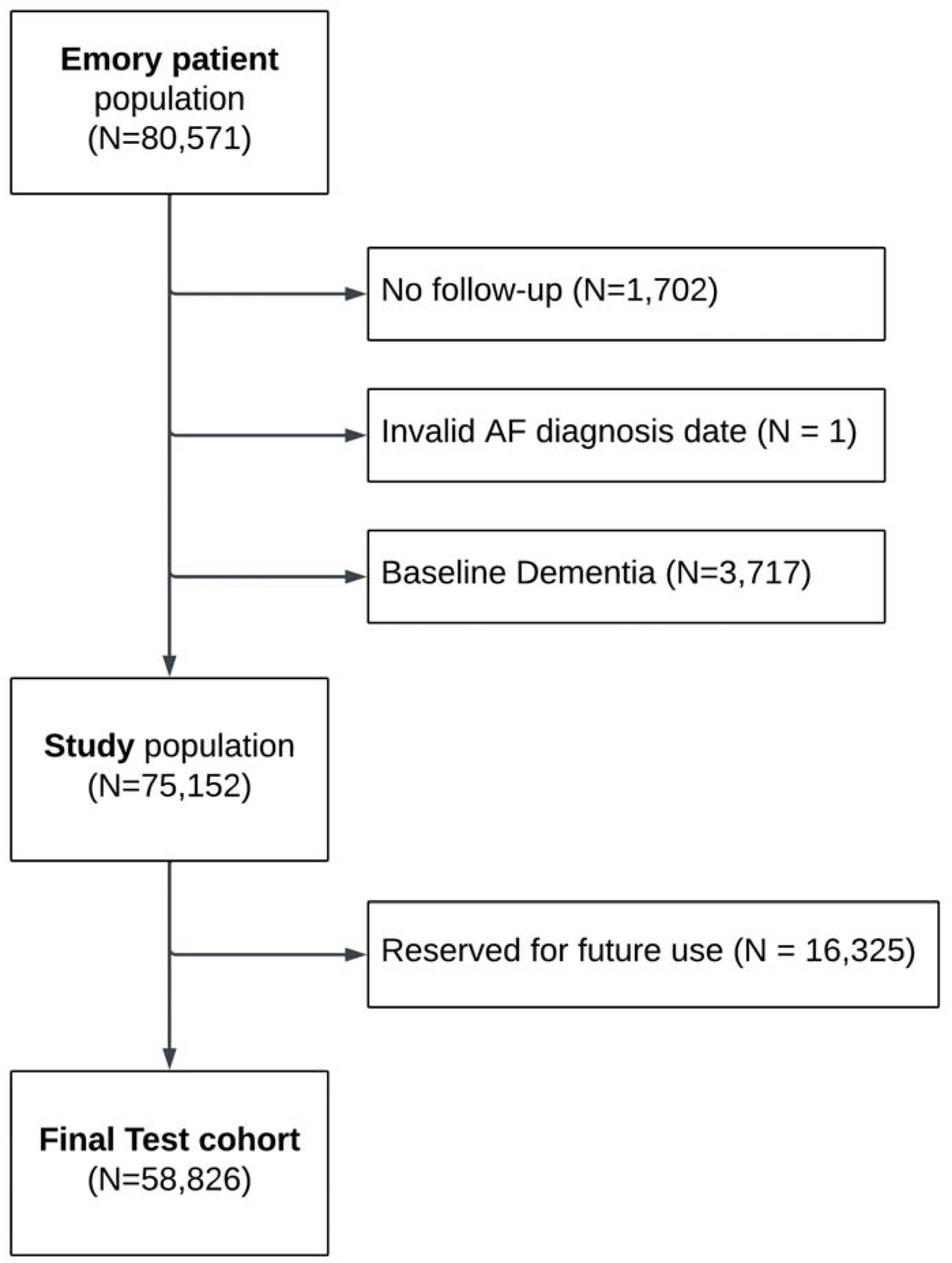
Consort Diagram for Emory external validation cohort.

**Extended Data Figure 2.**
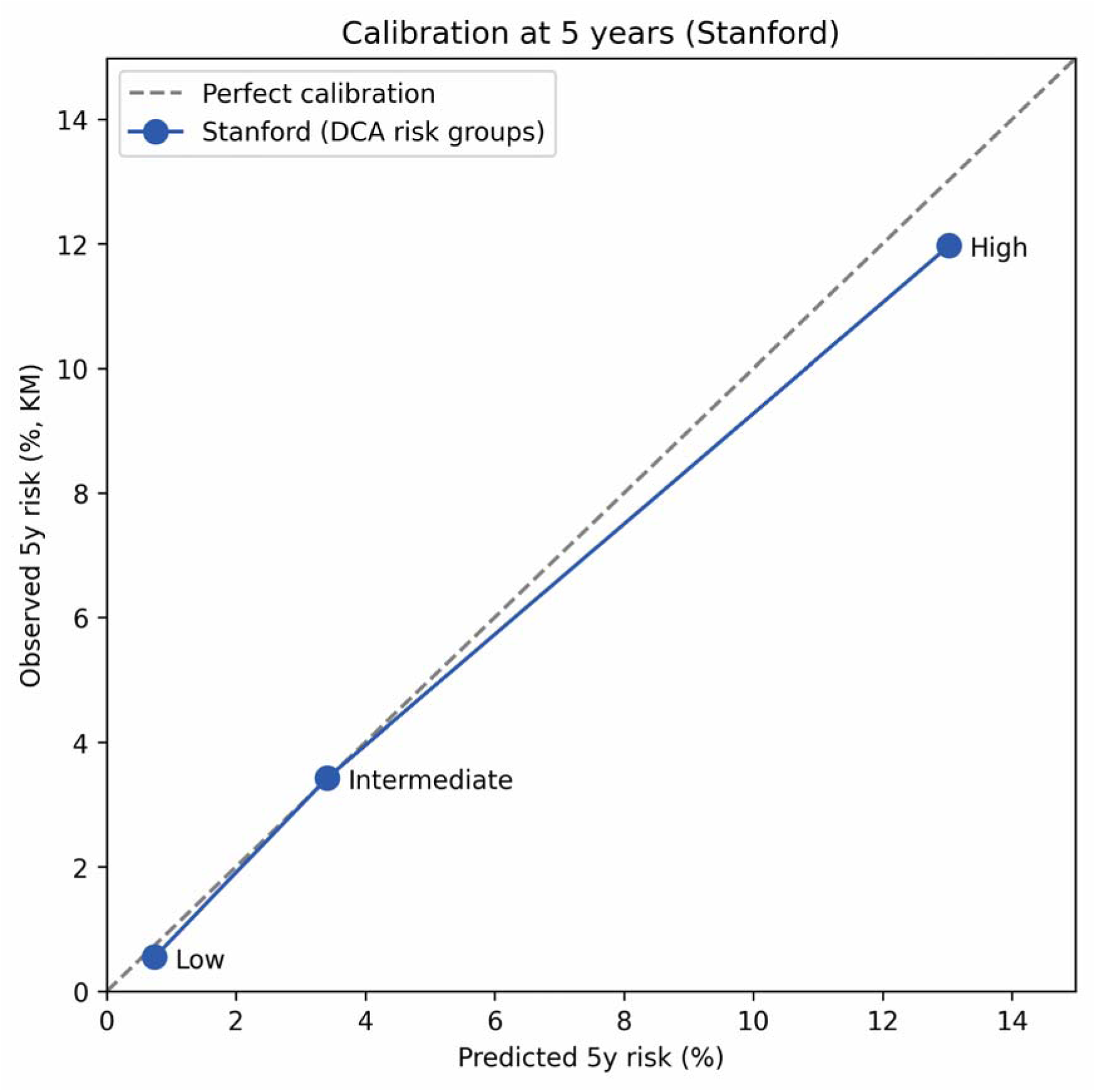
Observed versus Predicted 5 Year Dementia Incidence by Decision Curve Analysis Risk Group in Test Cohort. Risk groups were defined by 5-year predicted dementia risk thresholds derived from decision curve analysis. Predicted risk represents the mean model-estimated probability within each group. Observed risk was estimated by Kaplan–Meier analysis.

**Extended Data Figure 3.**
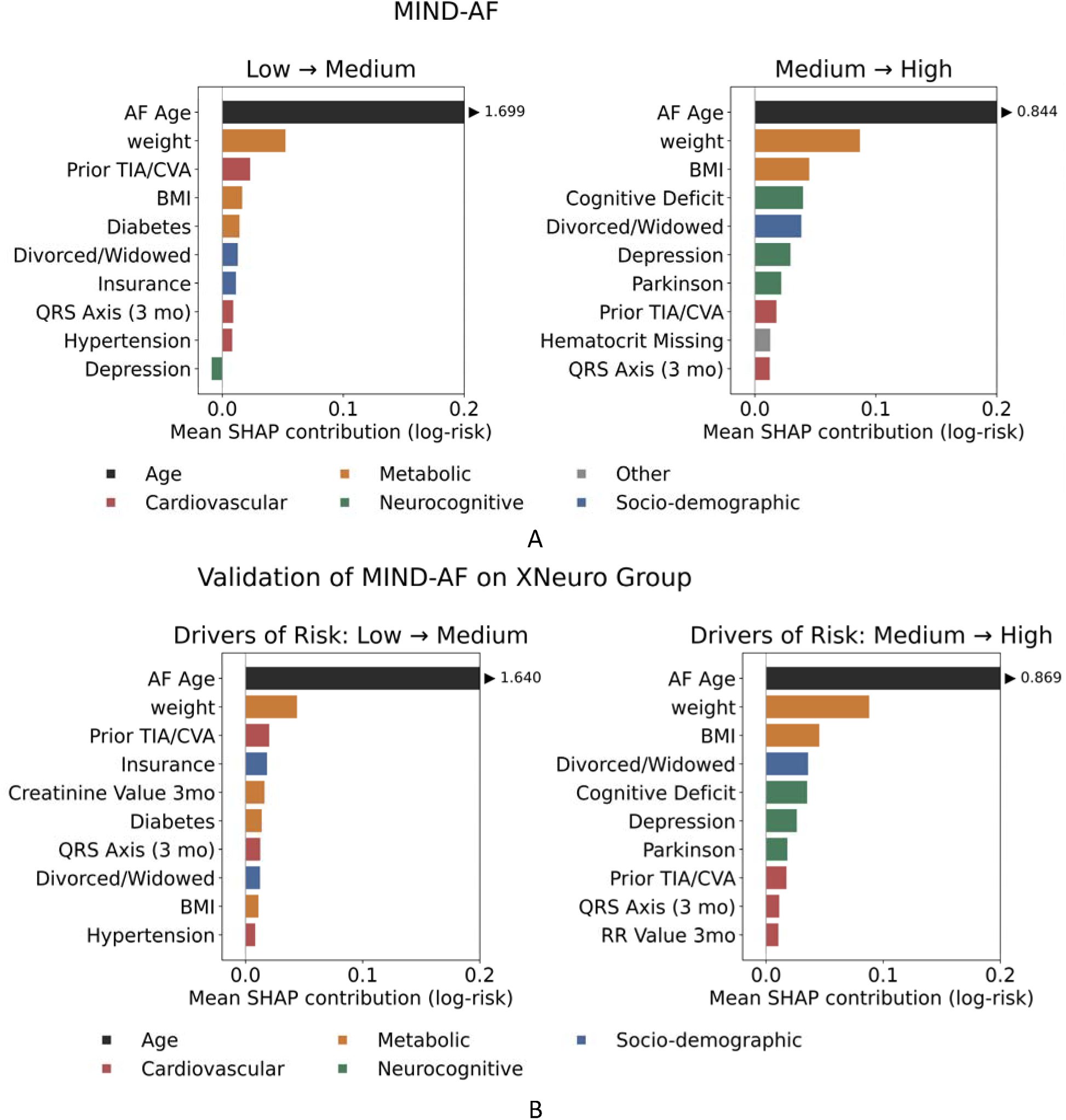
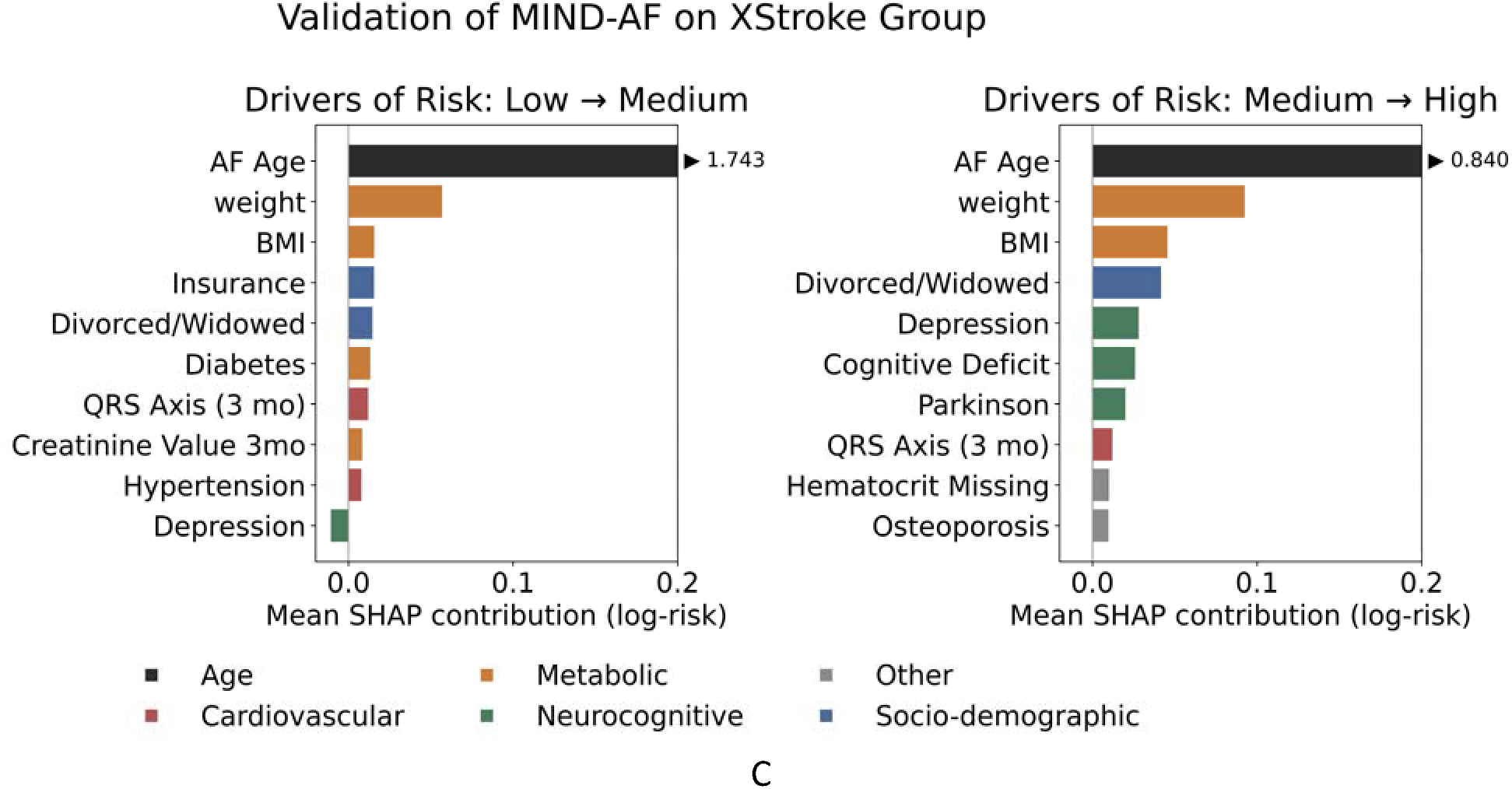
Top features driving risk between strata in MIND-AF. A. MIND-AF in the primary population excluding baseline dementia, B. MIND-AF in the X-neuro population where patients with baseline MCI are also excluded, C. MIND-AF in the X-stroke population where patients with baseline cerebrovascular disease are excluded. Features are ranked by SHAP importance, with colors denoting domains. Positive values indicate contribution to higher risk. The full feature set is provided in Supplementary Table 5.

**Extended Data Figure 4.**
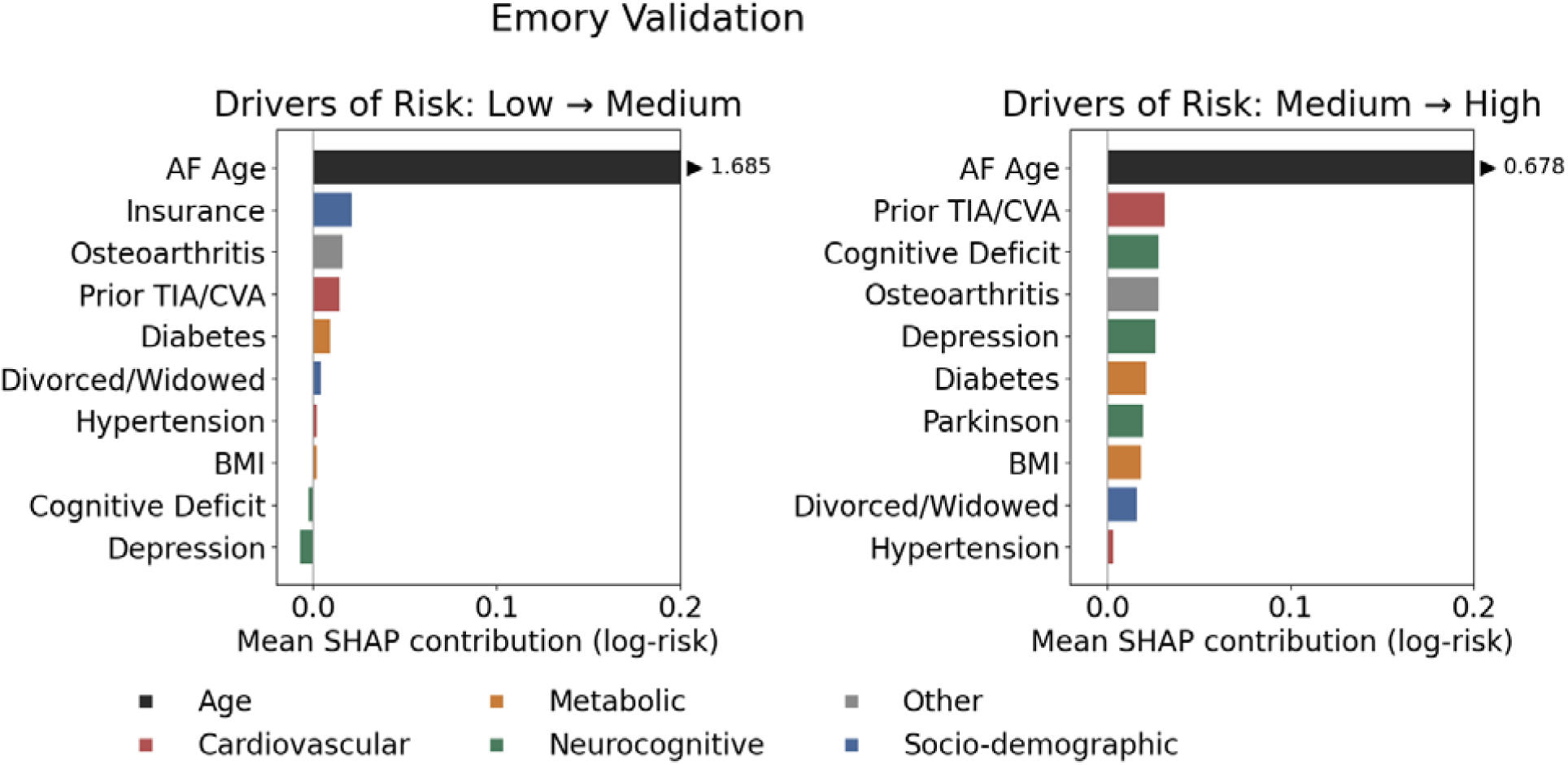
Top features driving risk between strata in MIND-AF in the external Georgia test cohort. Features are ranked by SHAP importance, with colors denoting domains. Positive values indicate contribution to higher risk.

**Extended Data Figure 5.**
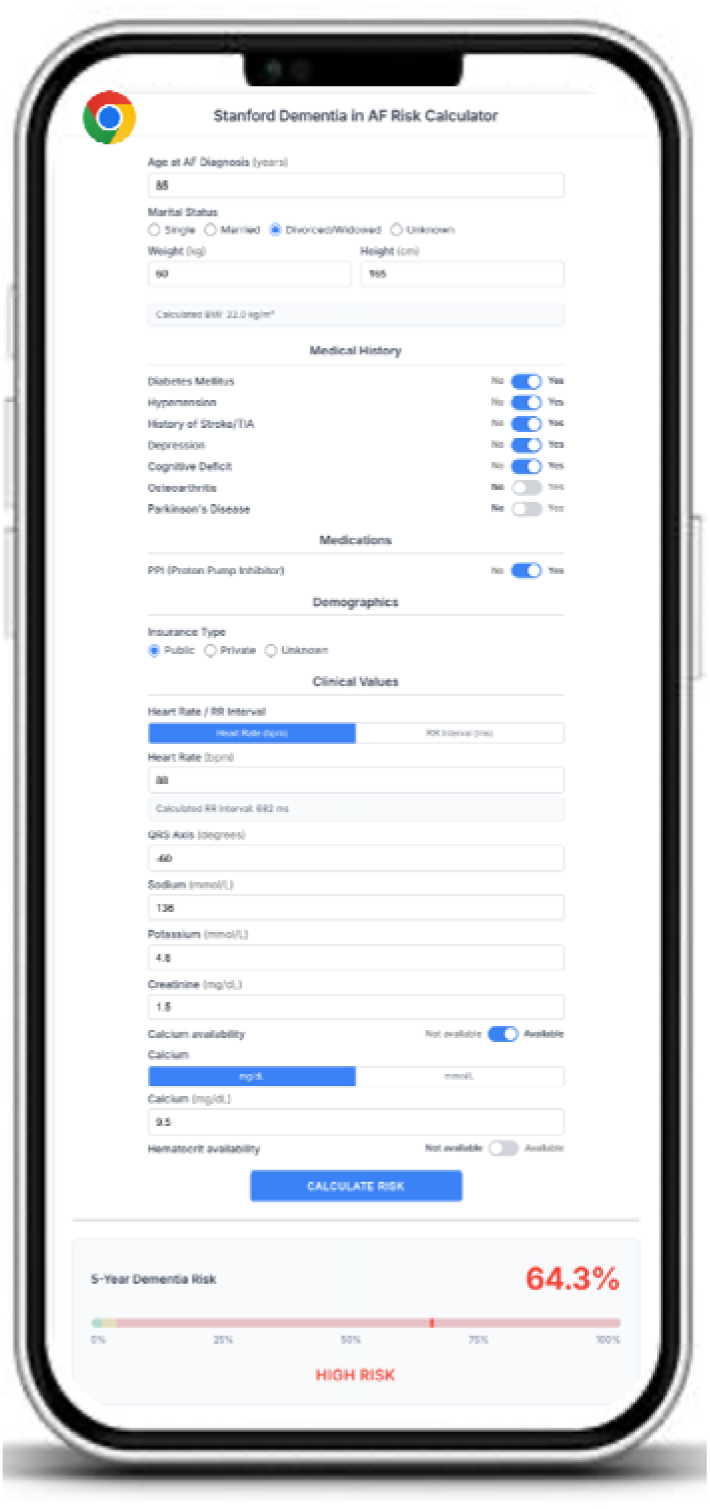
Clinical interface of the MIND-AF web application for 5-year dementia risk stratification in patients with atrial fibrillation. Representative output for a hypothetical 70-year-old male patient with AF, hypertension, history of stroke/TIA, and Parkinson’s disease, showing a predicted 5-year dementia risk of 64.3% (high risk). The calculator accepts 20 clinical features selected by BIC-guided ablation and returns a risk estimate with visual stratification into low, intermediate, and high-risk categories. The tool is available at https://github.com/NarayanLab/ml-dementia-prediction.

**Extended Data Table 1.** Comparison of different models in the MIND-AF validation set.

|  | **XGBoost** | **DeepSurv** | **GLMBoost** | **Cox-PH** |
|---|---|---|---|---|
| **C-index** | 0.870 | 0.861 | 0.844 | 0.847 |

Abbreviations. Cox-PH; Cox Proportional Hazards, DeepSurv; Deep Survival Model, GLM; Generalized Linear Models, XGBoost; Extreme Gradient Boosting

**Extended Data Table 2.** External validation performance (Emory University, Georgia) of MIND-AF versus Clinical Risk scores CHA₂DS₂-VASc and HATCH.

| Model | Time-to-dementia |  |  | Landmark Analysis (dementia within 5-years) |  |  |  |  |
|---|---|---|---|---|---|---|---|---|
|  | C-index (95% CI) | Hazard ratio low vs high (95% CI) | Hazard ratio per 1-SD (95% CI) | AUROC (95% CI) | Sensitivity (95% CI) | Specificity (95% CI) | Precision (95% CI) | F1-score (95% CI) |
| **Cohort without baseline dementia** |  |  |  |  |  |  |  |  |
| **MIND-AF** | 0.79 (0.78–0.80) | 11.06 (9.90–12.37) | 3.71 (3.51–3.92) | 0.83 (0.82–0.84) | 0.53 (0.50–0.56) | 0.89 (0.89–0.90) | 0.23 (0.22–0.25) | 0.32 (0.30–0.34) |
| *CHA<sub>2</sub>DS<sub>2</sub>-VASc* | 0.72 (0.71–0.73) | — | 2.22 (2.13–2.31) | 0.78 (0.77–0.79) | 0.90 (0.88–0.91) | 0.51 (0.50–0.51) | 0.10 (0.09–0.10) | 0.18 (0.17–0.19) |
| *HATCH* | 0.68 (0.67–0.69) | — | 1.84 (1.77–1.91) | 0.75 (0.74–0.76) | 0.47 (0.44–0.50) | 0.83 (0.83–0.84) | 0.14 (0.13–0.16) | 0.22 (0.21–0.24) |

Abbreviations. AF; Atrial fibrillation, AUROC; Area under receiver operating curve, C-index; Concordance index, MIND-AF; Modeling incident neurodegeneration in atrial fibrillation.

