## Supplemental Materials for "AI-Based Multi-Year Prediction of Incident Dementia in Patients with Atrial Fibrillation"

### Supplemental Methods

##### Competing Model Comparisons

ML models for this task included XGBoost, DeepSurv^1^, and GLMBoost^2^. The DeepSurv model was tuned using randomized Sobol search, as described in the original DeepSurv paper. We developed the hyperparameter grid in accordance with the WHAS model of the DeepSurv paper as this was the closest to our outcome of interest. Briefly, we implemented a feed-forward multi-layered perceptron model with 1-3 hidden layers, 32-96 units/layer, ReLU activations and dropout (0.05-0.30) ending in a linear log-risk node. We optimized the model using the Adam optimizer with exponential decay-based learning rate scheduling and gradient clipping. The initial learning rate was sampled log-uniformly between 1e-4–3e-3, and the loss was Cox partial likelihood. The model was trained to maximize the Harrell’s C-index in the dedicated validation set, following which the optimal model was refit on the training + validation set and evaluated on the held-out test set.

### Supplemental Results

#### Competing Model Comparisons

The best performing Cox PH model on the validation set was an unpenalized model (penalty = 0.0, L1 ratio = 0.0) with a C-index of 0.847 (95% CI 0.820-0.872). Refitting this model on a combined training + validation set yielded a C-index of 0.821 (95% CI 0.808-0.834) on the test set.

The DeepSurv model trained on the development set and applied to the validation set showed C-indices ranging from 0.805-0.845 across hyperparameter combinations. The best DeepSurv model had the following parameter combination: optimizer = adam, activation = relu, depth = 2, width = 79, dropout = 0.30, learning rate = 8e-4, L2 = 11.042, momentum = 0.873. This model when refit to the combined training + validation set yielded a C-index of 0.809 (95% CI 0.794-0.822) on the test set.

Other models were developed on the training dataset including Cox model with likelihood-based boosting and GLMBoost. However, these yielded inferior results compared to both XGBoost and DeepSurv. Furthermore, these models did not support GPU acceleration in python and hence were prohibitively slow during hyper-parameter tuning using randomized search.

#### Hyperparameters of Optimal Model (XGBoost)

The optimum performing XGBoost model had the following hyperparameter combination: max_depth = 3, min_child_weight = 3, gamma = 0.0, subsample = 0.8, colsample_bytree = 0.6, eta = 0.05, lambda = 0.0, alpha = 0.1, max_bin = 264 and had a validation C-index = 0.869 (95% CI 0.844-0.892).

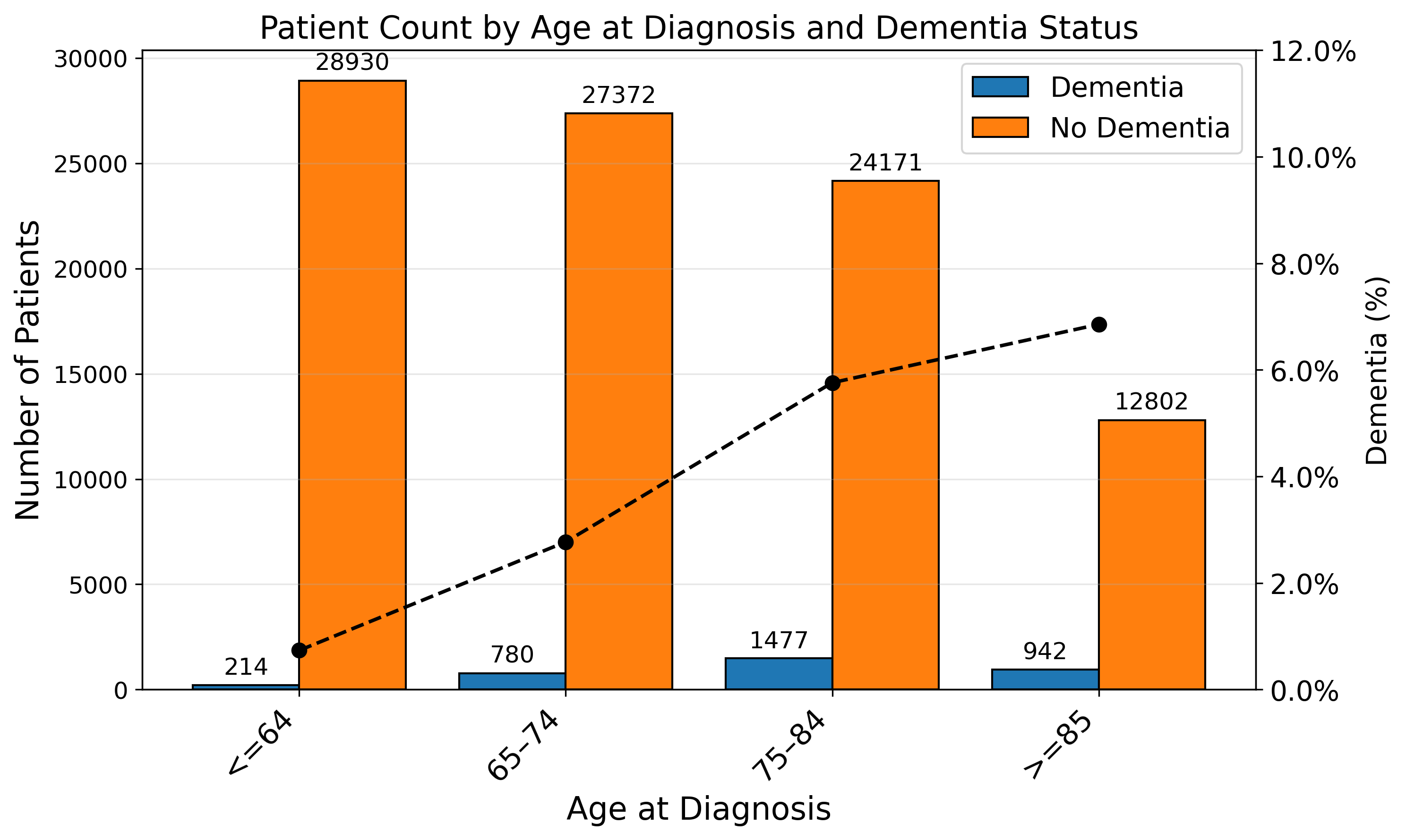

### Supplementary Figure 1. Dementia Status by Age of AF Diagnosis Age (Overall Cohort).

AF diagnosis age distribution of all patients within the primary MIND-AF cohort, for groups of ≤64 years, 65-74 years, 75-84 years, and ≥85 years color coded for presence (blue) and absence (orange) of dementia. The percentage of dementia in each age group is indicated on the dotted line corresponding to the right vertical axis.

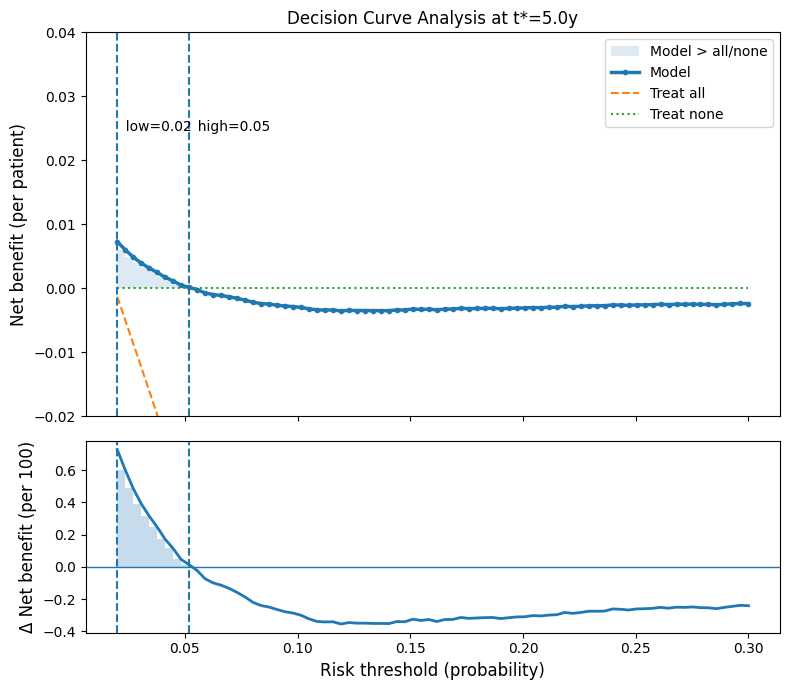

### Supplementary Figure 2. MIND-AF Decision Curve Analysis.

Decision curve analysis of all patients within the study population, calculated using the observed 5-year risk of dementia. The modeled decision curve crosses the X-Axis at 0.05.

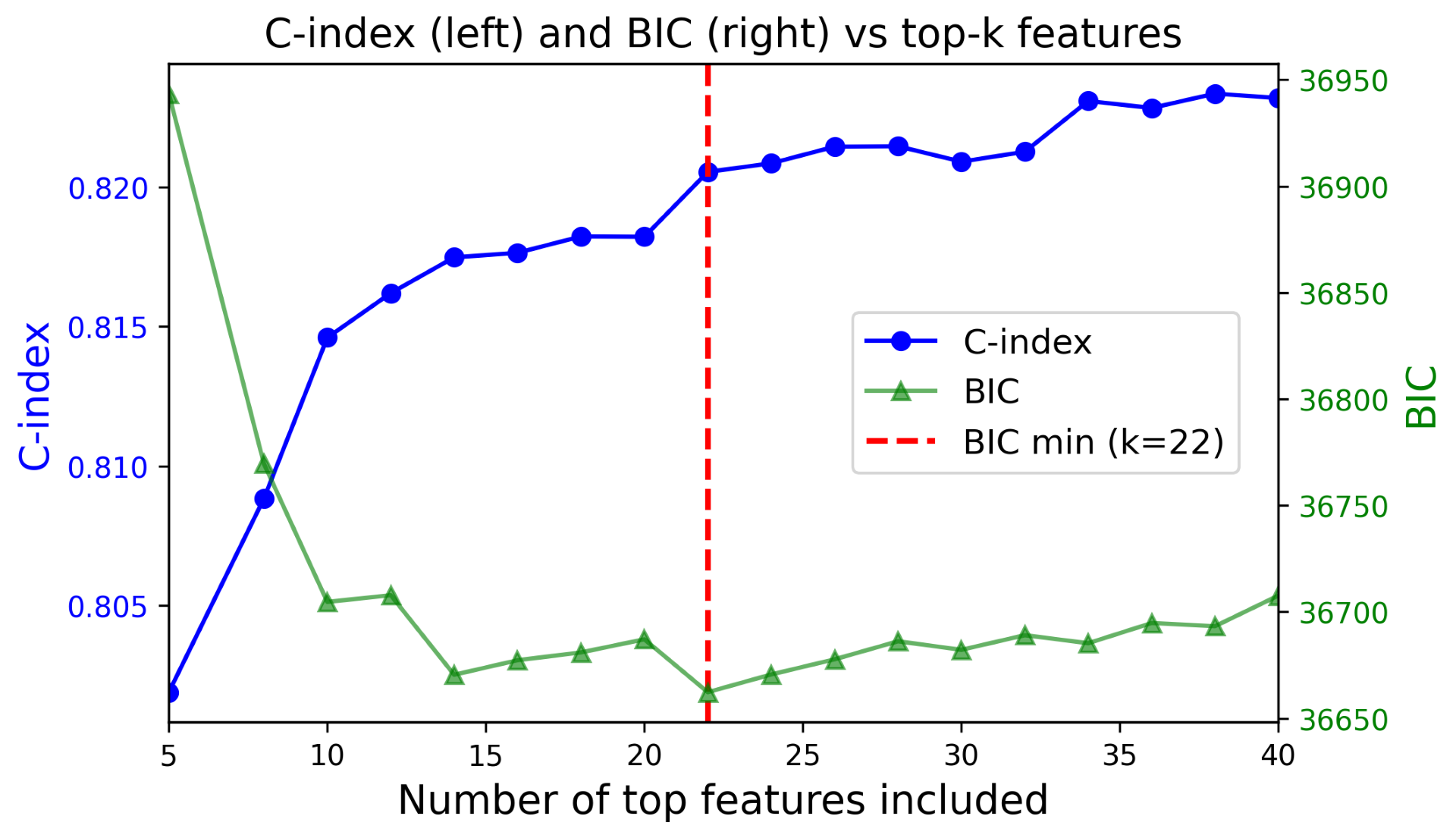

### Supplementary Figure 3. Selection of features for MIND-AF model hosted on web application.

Ablation analysis using Bayesian Information Criteria (BIC) selected the top 22 features as a minimal set did not significantly reduce performance of the original XGBoost model (DeLong’s test p=ns).

### Supplementary Table 1. Definitions for MIND-AF and ancillary models

| **Model** | **Category** | **Condition** | **ICD-10 codes** | **ICD-9 codes** |
| --- | --- | --- | --- | --- |
| **MIND-AF** | **Inclusion** | Atrial fibrillation | I48.0, I48.1, I48.2, I48.91 | 427.31 |
|  | **Exclusion** | Dementia | G30, F00, G31.83, F02, F01, G31.0, F03 | 294.10, 294.11, 294.20, 294.21, 331.0, 331.11, 331.19, 331.82, 290 |
|  | **Outcome** | Dementia | G30, F00, G31.83, F02, F01, G31.0, F03 | 294.10, 294.11, 294.20, 294.21, 331.0, 331.11, 331.19, 331.82, 290 |
| **MIND-AF-Xneuro** | **Inclusion** | Atrial fibrillation | I48.0, I48.1, I48.2, I48.91 | 427.31 |
|  | **Exclusion** | Dementia | G30, F00, G31.83, F02, G23.1, G31.85, G23.2, G23.3, G23.8, G90.3, F01, G31.0, F03 | 294.10, 294.11, 294.20, 294.21, 331.0, 331.11, 331.19, 331.82, 290 |
|  |  | Mild cognitive impairment | G31.84, F06.7 | 331.83 |
|  |  | ALS-FTD spectrum / neurodegenerative | G12.21–G12.25, G10, A81.0, A81.81, A81.82, A81.83, A81.9 | — |
|  | **Outcome** | Dementia | G30, F00, G31.83, F02, G23.1, G31.85, G23.2, G23.3, G23.8, G90.3, F01, G31.0, F03 | 294.10, 294.11, 294.20, 294.21, 331.0, 331.11, 331.19, 331.82, 290 |
| **MIND-AF-Xstroke** | **Inclusion** | Atrial fibrillation | I48.0, I48.1, I48.2, I48.91 | 427.31 |
|  | **Exclusion** | Dementia | G30, F00, G31.83, F02, F01, G31.0, F03 | 294.10, 294.11, 294.20, 294.21, 331.0, 331.11, 331.19, 331.82, 290 |
|  |  | Stroke / TIA | I60, I61, I62, I63, I64, I65, I66, G45, G46 | 435, 436, 438, 433.01, 433.21, 433.31, 433.81, 433.91, 434.01, 434.11, 434.91 |
|  | **Outcome** | Dementia | G30, F00, G31.83, F02, F01, G31.0, F03 | 294.10, 294.11, 294.20, 294.21, 331.0, 331.11, 331.19, 331.82, 290 |

Abbreviations. AF; Atrial fibrillation, ALS; Amyotrophic Lateral Sclerosis, MCI; Mild Cognitive Impairment, MIND-AF; Modeling Incident Neurodegeneration in AF; TIA; Transient Ischemic Attack.

### Supplementary Table 2. Complete characteristics of the California MIND-AF population.

|  | **MIND-AF study population (Stanford)** | | |
| --- | --- | --- | --- |
| **Feature** | **No dementia (n = 93,275)** | **Dementia (n = 3,413)** | **P value** |
| **Demographics** | | | |
| AF diagnosis age, years | 70.2 ± 14.1 | 79.2 ± 9.1 | **<0.001** |
| Age at last follow-up, years | 74.0 ± 13.9 | 87.0 ± 8.1 | **<0.001** |
| BMI, kg/m² | 27.3 ± 5.5 | 25.0 ± 5.0 | **<0.001** |
| Weight, kg | 79.7 ± 19.4 | 69.7 ± 16.5 | **<0.001** |
| Height, cm | 170.1 ± 10.3 | 166.6 ± 10.5 | **<0.001** |
| Female sex, n (%) | 38,925 (41.7) | 1,815 (53.2) | **<0.001** |
| **Race and ethnicity** | | | |
| White, n (%) | 60,417 (64.8) | 2,452 (71.8) | **<0.001** |
| Hispanic or Latino, n (%) | 7,006 (7.5) | 184 (5.4) | **<0.001** |
| **Social characteristics** | | | |
| Married, n (%) | 55,420 (59.4) | 1,635 (47.9) | **<0.001** |
| Private insurance, n (%) | 53,825 (57.7) | 2,444 (71.6) | **<0.001** |
| Never smoker, n (%) | 48,457 (52.0) | 1,907 (55.9) | **<0.001** |
| Area Deprivation Index – National Rank | 7.6 ± 10.2 | 7.8 ± 10.4 | 0.497 |
| **Cardiovascular comorbidities** | | | |
| Congestive heart failure, n (%) | 19,586 (21.0) | 607 (17.8) | **<0.001** |
| Hypertension, n (%) | 50,956 (54.6) | 2,180 (63.9) | **<0.001** |
| Hyperlipidaemia, n (%) | 37,549 (40.3) | 1,554 (45.5) | **<0.001** |
| Stroke or TIA, n (%) | 11,450 (12.3) | 580 (17.0) | **<0.001** |
| Coronary artery disease, n (%) | 26,136 (28.0) | 1,011 (29.6) | **0.041** |
| Myocardial infarction, n (%) | 4,975 (5.3) | 197 (5.8) | 0.264 |
| Ventricular tachycardia, n (%) | 5,546 (5.9) | 150 (4.4) | **<0.001** |
| Peripheral artery disease, n (%) | 4,717 (5.1) | 233 (6.8) | **<0.001** |
| COPD, n (%) | 12,907 (13.8) | 513 (15.0) | **0.048** |
| Obstructive sleep apnea, n (%) | 10,815 (11.6) | 316 (9.3) | **<0.001** |
| LVEF | 58.0 ± 6.6 | 58.6 ± 4.1 | **<0.001** |
| **Non-cardiovascular comorbidities** | | | |
| Diabetes mellitus, n (%) | 16,832 (18.0) | 654 (19.2) | 0.096 |
| Chronic kidney disease, n (%) | 14,052 (15.1) | 477 (14.0) | 0.080 |
| Asthma, n (%) | 6,885 (7.4) | 273 (8.0) | 0.176 |
| Osteoarthritis, n (%) | 10,794 (11.6) | 597 (17.5) | **<0.001** |
| Osteoporosis, n (%) | 4,526 (4.9) | 338 (9.9) | **<0.001** |
| Rheumatoid arthritis, n (%) | 1,501 (1.6) | 88 (2.6) | **<0.001** |
| Cancer, n (%) | 15,492 (16.6) | 507 (14.9) | **0.007** |
| Depression, n (%) | 8,159 (8.7) | 443 (13.0) | **<0.001** |
| Epilepsy, n (%) | 1,391 (1.5) | 51 (1.5) | 0.989 |
| Parkinson's disease, n (%) | 727 (0.8) | 88 (2.6) | **<0.001** |
| Schizophrenia, n (%) | 685 (0.7) | 39 (1.1) | **0.007** |
| Thyroid disorders, n (%) | 12,558 (13.5) | 639 (18.7) | **<0.001** |
| Hearing deficiency, n (%) | 5,066 (5.4) | 280 (8.2) | **<0.001** |
| Vision impairment, n (%) | 924 (1.0) | 59 (1.7) | **<0.001** |
| Brain injury, n (%) | 867 (0.9) | 48 (1.4) | **0.005** |
| Cognitive deficit, n (%) | 3,073 (3.3) | 237 (6.9) | **<0.001** |
| Mild cognitive impairment, n (%) | 462 (0.5) | 68 (2.0) | **<0.001** |
| **Laboratory values** | | | |
| Sodium, mmol/l | 138.6 ± 2.8 | 138.4 ± 2.9 | **0.003** |
| Potassium, mmol/l | 4.2 ± 0.4 | 4.2 ± 0.4 | **<0.001** |
| Creatinine, mg/dl | 1.1 ± 0.7 | 1.1 ± 0.5 | **<0.001** |
| Calcium, mg/dl | 8.6 ± 1.8 | 8.7 ± 1.6 | **0.001** |
| INR | 1.3 ± 0.4 | 1.3 ± 0.5 | **<0.001** |
| **ECG values** | | | |
| RR interval, ms | 780.0 ± 151.1 | 774.8 ± 130.2 | **0.023** |
| QTc interval, ms | 434.7 ± 26.5 | 433.7 ± 22.5 | **0.011** |
| QRS axis, degrees | 16.9 ± 39.2 | 12.5 ± 33.5 | **<0.001** |
| **Medications** | | | |
| Aspirin, n (%) | 24,076 (25.8) | 861 (25.2) | 0.443 |
| NOAC, n (%) | 10,034 (10.8) | 179 (5.2) | **<0.001** |
| Coumadin/Warfarin, n (%) | 6,656 (7.1) | 254 (7.4) | 0.495 |
| HMG-CoA reductase inhibitors, n (%) | 29,431 (31.6) | 891 (26.1) | **<0.001** |
| ACE inhibitor or ARB, n (%) | 24,410 (26.2) | 815 (23.9) | **0.003** |
| Beta-blockers, n (%) | 28,771 (30.8) | 853 (25.0) | **<0.001** |
| Amiodarone, n (%) | 3,702 (4.0) | 72 (2.1) | **<0.001** |
| **Procedures** | | | |
| CABG, n (%) | 423 (0.5) | 38 (1.1) | **<0.001** |
| Valvular surgery, n (%) | 2,576 (2.8) | 63 (1.8) | **0.001** |
| Catheter ablation for AF, n (%) | 198 (0.2) | 0 (0.0) | **0.007** |

*Bold P values indicate statistical significance (P < 0.05). AF, atrial fibrillation; TIA, transient ischemic attack; COPD, chronic obstructive pulmonary disease; LVEF, left ventricular ejection fraction; INR, international normalized ratio; NOAC, novel oral anticoagulant; ACE, angiotensin-converting enzyme; ARB, angiotensin receptor blocker; CABG, coronary artery bypass graft; HMG-CoA, 3-hydroxy-3-methylglutaryl-coenzyme A.*

### Supplementary Table 3. ICD-9 and ICD-10 Codes used to define Comorbid Conditions

| **Condition** | **Coding system** | **Codes** |
| --- | --- | --- |
| **Atrial Flutter** | ICD-10 | I48.3, I48.4, I48.92 |
|  | ICD-9 | 427.32 |
| **Congestive Heart Failure** | ICD-10 | I50.X |
|  | ICD-9 | 428.X |
| **Hypertension** | ICD-10 | I10, I11.X, I12.X, I13.X, I15.X |
|  | ICD-9 | 401.X, 402.X, 403.X, 404.X, 405.X |
| **Hyperlipidemia** | ICD-10 | E78.0X, E78.1, E78.2, E78.3, E78.4X, E78.5 |
|  | ICD-9 | 272.0, 272.1, 272.2, 272.3, 272.4 |
| **Coronary Artery Disease** | ICD-10 | I20.X, I21.X, I22.X, I23.X, I24.X, I25.X |
|  | ICD-9 | 410.X, 411.X, 412, 413.X, 414.X |
| **Myocardial Infarction** | ICD-10 | I21.X |
|  | ICD-9 | 410.X |
| **Ventricular Tachycardia** | ICD-10 | I47.2X |
|  | ICD-9 | 427.1 |
| **Peripheral Artery Disease** | ICD-10 | I73.1, I73.8X, I73.9, I74.3, I74.4, I74.5 |
|  | ICD-9 | 440.2X, 440.3X, 440.8, 440.9, 443.9 |
| **COPD** | ICD-10 | J40, J41.X, J42, J43.X, J44.X |
|  | ICD-9 | 490, 491, 492, 493.X, 494.X, 495.X, 496 |
| **Obstructive Sleep Apnea** | ICD-10 | G47.30, G47.33 |
|  | ICD-9 | 327.23, 780.51, 780.53, 780.57 |
| **Diabetes Mellitus** | ICD-10 | E08.X, E09.X, E10.X, E11.X, E13.X |
|  | ICD-9 | 250.X |
| **Chronic Kidney Disease** | ICD-10 | N18.X |
|  | ICD-9 | 585.X |
| **Asthma** | ICD-10 | J45.X |
|  | ICD-9 | 493.X |
| **Osteoarthritis** | ICD-10 | M15.X, M16.X, M17.X, M18.X, M19.X |
|  | ICD-9 | 715.00, 715.1X |
| **Osteoporosis** | ICD-10 | M80.X, M81.X |
|  | ICD-9 | 733.0X |
| **Rheumatoid Arthritis** | ICD-10 | M05.X, M06.X |
|  | ICD-9 | 714.0 |
| **Cancer** | ICD-10 | C00.X–C97.X |
|  | ICD-9 | 140.X, 172.X, 174.X, 195.X–208.X, 238.6 |
| **Depression** | ICD-10 | F31.3X, F31.4, F31.5, F31.6X, F32.0–F32.9, F33.0–F33.8, F34.1, F41.2 |
|  | ICD-9 | 296.20, 296.25, 296.30, 296.35, 296.5, 296.6, 296.82, 300.4, 309.0, 309.1, 309.28, 311 |
| **Epilepsy** | ICD-10 | G40.X, G41.X |
|  | ICD-9 | 345.X |
| **HIV/AIDS** | ICD-10 | B20.X, B21.X, B22.X, B24 |
|  | ICD-9 | 042 |
| **Parkinson's Disease** | ICD-10 | F02.3, G20.X |
|  | ICD-9 | 332.0 |
| **Schizophrenia** | ICD-10 | F20.X, F21.X, F22.X, F25.X, F28.X, F29.X |
|  | ICD-9 | 295.X |
| **Tuberculosis** | ICD-10 | A15.X |
|  | ICD-9 | 010.X, 011.X, 012.X |
| **Vision Impairment** | ICD-10 | H54.0X, H54.1X, H54.2X, H54.3, H54.8 |
|  | ICD-9 | 369.X |
| **Cognitive Impairment** | ICD-10 | F06.7, R41.3, R41.8 |
|  | ICD-9 | 331.83 |
| **Brain Injury** | ICD-10 | S06.X |
|  | ICD-9 | 850.X, 851.X, 852.X, 853.X, 854.X |
| **Alcohol Abuse** | ICD-10 | F10.0, F10.2, F10.3 |
|  | ICD-9 | 303.X |
| **Thyroid Disorders** | ICD-10 | E03.5, E03.8, E03.9, E05.0–E05.9, E06.2 |
|  | ICD-9 | 240.X, 241.X, 242.X, 244.X, 245.X, 246.X |
| **Hearing Deficiency** | ICD-10 | H90.X |
|  | ICD-9 | 389.X |
| **Vitamin B12 Deficiency** | ICD-10 | D51.X |
|  | ICD-9 | 281.1 |
| **Lupus** | ICD-10 | M32.X |
|  | ICD-9 | 710.0 |

### Supplementary Table 4. Risk groups by Decision Curve Analysis in the MIND-AF model applied to different cohort definitions

| **Model** | **Low risk** | **Intermediate risk** | **High risk** |
| --- | --- | --- | --- |
|  | *(<2%)* | *(2–5.2% or 2–4.8%)* | *(>5.2% or >4.8%)* |
| **MIND-AF on original cohort** | **< 2%** (n = 14,461) | **2–5.2%** (n = 7,891) | **> 5.2%** (n = 9,331) |
| **MIND-AF on Xneuro cohort** | **< 2%** (n = 14,043) | **2–4.8%** (n = 7,219) | **> 4.8%** (n = 10,345) |
| **MIND-AF on Xstroke cohort** | **< 2%** (n = 13,092) | **2–5.2%** (n = 6,587) | **> 5.2%** (n = 6,832) |

Abbreviations. AF; Atrial Fibrillation, DCA; Decision Curve Analysis.

### Supplementary Table 5. Explainability Featured (mean SHAP values) of top 25 clinical features by DCA-defined risk group in the MIND-AF model

| **Clinical feature** | **Category** | **Low risk** | **Intermediate risk** | **High risk** |
| --- | --- | --- | --- | --- |
|  |  | (<2%) | (2–5.2%) | (>5.2%) |
| AF diagnosis age, years | *Demographics* | -1.1633 | 0.5353 | 1.3792 |
| Body weight, kg | *Demographics* | -0.0447 | 0.0076 | 0.0944 |
| Body mass index, kg/m² | *Demographics* | -0.0192 | -0.0028 | 0.0422 |
| Marital status | *Demographics* | -0.0112 | 0.0017 | 0.0401 |
| Depression | *Comorbidities* | 0.0051 | -0.0037 | 0.0257 |
| TIA/stroke/CVA | *Comorbidities* | -0.0169 | 0.0062 | 0.0240 |
| Cognitive deficit | *Comorbidities* | -0.0234 | -0.0221 | 0.0178 |
| Diabetes mellitus | *Comorbidities* | 0.0016 | 0.0159 | 0.0208 |
| RR interval, ms | *ECG* | -0.0170 | -0.0130 | -0.0031 |
| QRS axis, degrees | *ECG* | -0.0062 | 0.0029 | 0.0153 |
| Insurance type | *Socioeconomic* | 0.0028 | 0.0141 | 0.0123 |
| Creatinine, mg/dL | *Laboratory* | 0.0024 | 0.0100 | 0.0138 |
| Osteoarthritis | *Comorbidities* | -0.0130 | -0.0055 | 0.0037 |
| Parkinson's disease | *Comorbidities* | -0.0094 | -0.0092 | 0.0126 |
| Propafenone | *Medications* | 0.0039 | 0.0082 | 0.0110 |
| Hematocrit (missing) | *Laboratory* | -0.0013 | -0.0018 | 0.0109 |
| Osteoporosis | *Comorbidities* | -0.0097 | -0.0064 | 0.0033 |
| Serum calcium, mg/dL | *Laboratory* | -0.0089 | -0.0045 | -0.0014 |
| Serum sodium, mmol/L | *Laboratory* | -0.0083 | -0.0055 | 0.0029 |
| Hypertension | *Comorbidities* | -0.0075 | 0.0007 | 0.0081 |
| Mild cognitive impairment | *Comorbidities* | -0.0077 | -0.0073 | 0.0033 |
| Chronic kidney disease | *Comorbidities* | 0.0014 | 0.0042 | 0.0073 |
| Serum calcium (missing) | *Laboratory* | -0.0013 | -0.0020 | 0.0060 |
| Height, cm | *Demographics* | 0.0060 | 0.0060 | 0.0021 |
| QTc interval, ms | *ECG* | 0.0027 | 0.0027 | 0.0053 |

Abbreviations. AF; Atrial fibrillation, QTc, corrected QT interval; RR, R–R interval; TIA; Transient Ischemic attack.

P.S. Positive SHAP values indicate that the feature increases predicted dementia risk; negative values indicate a risk-reducing contribution. Features are ranked by the maximum absolute SHAP value across all three risk groups. Risk groups were defined by 5-year predicted dementia risk thresholds derived from decision curve analysis: low (<2%), intermediate (2–5.2%), and high (>5.2%). SHAP values represent mean contributions across all patients within each group. Only the top 25 of n = 91 model features are shown; remaining features had negligible contributions (mean |SHAP| <0.001 across all groups).

### Supplementary Table 6. Medication Curation by String Search

| **Medication class** | **Search strings** |
| --- | --- |
| **Aspirin** | Aspirin |
| **P2Y12 Inhibitors** | Clopidogrel, Ticagrelor, Prasugrel |
| **Direct Oral Anticoagulants (DOAC)** | Apixaban, Dabigatran, Edoxaban |
| **Warfarin** | Warfarin, Coumadin |
| **HMG-CoA Reductase Inhibitors** | Rosuvastatin, Atorvastatin, Pravastatin, Simvastatin, Fluvastatin, Pitavastatin, Lovastatin |
| **ACE Inhibitors / ARBs** | Telmisartan, Lisinopril, Benazepril, Olmesartan, Valsartan, Irbesartan, Losartan, Quinapril, Captopril, Ramipril, Azilsartan, Fosinopril, Enalapril, Candesartan, Perindopril |
| **Beta-Blockers** | Sotalol, Metoprolol, Carvedilol, Propranolol, Nebivolol, Atenolol, Timolol, Bisoprolol, Pindolol, Nadolol, Acebutolol |
| **Low Molecular Weight Heparin** | Enoxaparin |
| **GDMT** | Eplerenone, Spironolactone, Metoprolol, Carvedilol, Dapagliflozin, Nebivolol, Valsartan, Bisoprolol |
| **Calcium Channel Blockers** | Diltiazem, Verapamil |
| **Flecainide** | Flecainide |
| **Digoxin** | Digoxin |
| **Propafenone** | Propafenone |
| **Amiodarone** | Amiodarone |
| **Dronedarone** | Dronedarone |
| **Sotalol** | Sotalol |
| **Dofetilide** | Dofetilide |
| **Anticholinergics** | Tolterodine, Promethazine, Olanzapine, Hyoscyamine, Scopolamine, Diphenhydramine, Benztropine, Carisoprodol, Atropine, Cyclobenzaprine, Glycopyrrolate, Dicyclomine, Oxybutynin, Methocarbamol, Amitriptyline, Trospium, Solifenacin, Orphenadrine, Chlorpheniramine, Doxepin, Hydroxyzine, Nortriptyline, Clomipramine, Homatropine, Imipramine, Fesoterodine, Darifenacin, Trihexyphenidyl, Propantheline, Cyproheptadine, Perphenazine, Clozapine, Brompheniramine |
| **Sedatives** | Ramelteon, Clonazepam, Zolpidem, Lorazepam, Phenobarbital, Temazepam, Diazepam, Midazolam, Eszopiclone, Alprazolam, Flurazepam, Oxazepam, Zaleplon, Secobarbital |
| **Proton Pump Inhibitors** | Pantoprazole, Esomeprazole, Omeprazole, Lansoprazole, Rabeprazole, Dexlansoprazole |
| **Antipsychotics** | Quetiapine, Olanzapine, Lurasidone, Haloperidol, Aripiprazole, Ziprasidone, Risperidone |
| **Opioids** | Fentanyl, Morphine, Codeine, Hydrocodone, Hydromorphone, Oxycodone, Methadone, Buprenorphine, Tramadol, Oxymorphone |
| **GLP-1 Agonists** | Semaglutide, Exenatide, Dulaglutide, Liraglutide |
| **Glucocorticoids** | Methylprednisolone, Dexamethasone, Prednisone, Hydrocortisone, Budesonide |

**Abbreviations:** ACE, angiotensin-converting enzyme; ARB, angiotensin receptor blocker; DOAC, direct oral anticoagulant; GDMT, guideline-directed medical therapy; GLP-1, glucagon-like peptide-1; HMG-CoA, hydroxymethylglutaryl-coenzyme A.
